# Express yourself: Quantitative real-time PCR assays for rapid chromosomal antimicrobial resistance detection in *Pseudomonas aeruginosa*

**DOI:** 10.1101/2022.02.03.22270419

**Authors:** Danielle E. Madden, Olusola Olagoke, Timothy Baird, Jane Neill, Kay A. Ramsay, Tamieka A. Fraser, Scott C. Bell, Derek S. Sarovich, Erin P. Price

**Author notes:** Corresponding authors: Derek Sarovich, Erin Price. These authors contributed equally (Derek S. Sarovich, Erin P. Price).

## Abstract

The rise of antimicrobial-resistant (AMR) bacteria is a global health emergency. One critical facet in tackling this epidemic is more rapid AMR diagnosis in serious multi-drug resistant pathogens like *Pseudomonas aeruginosa*. Here, we designed and then validated two multiplex quantitative real-time PCR (qPCR) assays to simultaneously detect differential expression of the resistance-nodulation-division efflux pumps MexAB-OprM, MexCD-OprJ, MexEF-OprN, MexXY-OprM, the AmpC β-lactamase, and the porin OprD, which are commonly associated with chromosomally-encoded AMR. Next, qPCRs were tested on 15 sputa from 11 participants with *P. aeruginosa* respiratory infections to determine AMR profiles *in vivo*. We confirm multiplex qPCR testing feasibility directly on sputa, representing a key advancement in *in vivo* AMR diagnosis. Notably, comparison of sputa with their derived isolates grown in Luria-Bertani broth (±2.5% NaCl) or a 5-antibiotic cocktail showed marked expression differences, illustrating the difficulty in replicating *in vivo* expression profiles *in vitro*. Cystic fibrosis sputa showed significantly reduced *mexE* and *mexY* expression when compared with chronic obstructive pulmonary disease sputa, despite harbouring fluoroquinolone- and aminoglycoside-resistant strains, indicating that these loci are not contributing to AMR *in vivo. oprD* was also significantly downregulated in cystic fibrosis sputa, even in the absence of contemporaneous carbapenem use, suggesting a common adaptive trait in chronic infections that may affect carbapenem efficacy. Sputum *ampC* expression was highest in participants receiving carbapenems (6.7-15x), some of whom were simultaneously receiving cephalosporins, the latter of which would be rendered ineffective by the upregulated *ampC*. Our qPCR assays provide valuable insights into the *P. aeruginosa* resistome, and their use on clinical specimens will permit timely treatment alterations that will improve patient outcomes and antimicrobial stewardship measures.

## Introduction

The persistent rise in the prevalence and dissemination of antimicrobial-resistant (AMR) bacteria threatens to push humankind towards a post-antibiotic world (1) where untreatable superbugs endanger global health and food security and cause significant morbidity, mortality and economic burden (2). The opportunistic Gram-negative bacterium, *Pseudomonas aeruginosa*, is a member of the ‘ESKAPE’ group of AMR pathogens that represent the greatest concern to global health (3). *P. aeruginosa* can cause life-threatening infections (4), including in those with chronic respiratory diseases where localised immune defences are compromised such as cystic fibrosis (CF) or chronic obstructive pulmonary disease (COPD) (5). Once infection is established, *P. aeruginosa* eradication is made more difficult by many factors, including its high intrinsic AMR and its capacity to develop multi-drug resistance (MDR) to all classes of clinically-relevant antibiotics (6).

AMR diagnostics are a critical weapon in our battle against superbugs as they enable rapid, cost-effective, and high-throughput AMR detection, leading to more judicious and targeted antibiotic use (7). However, a lack of such diagnostics means that broad-spectrum antimicrobials are commonly used in the clinic in lieu of personalised treatments guided by antimicrobial sensitivity, genotype, and gene expression data (8). This practice, exacerbated during the COVID-19 pandemic (9), has unfortunately contributed to the emergence, persistence, and spread of AMR and MDR pathogens worldwide, with potentially devastating outcomes including treatment failure and mortality (10). Advancements in next-generation sequencing (NGS) technologies are enabling close-to-real-time identification of AMR (11). However, it is currently impractical and costly to use these technologies for rapid, high-throughput detection (12); as such, their implementation in routine clinical diagnostics is not yet feasible. Instead, nucleic acid-based detection platforms such as real-time PCR remain vital – and substantially cheaper, simpler, and more accessible in routine clinical microbiology services than NGS – for the rapid identification of pathogens and their AMR determinants (12). In addition to identifying AMR-conferring single-nucleotide polymorphisms [SNPs] (13, 14), insertions/deletions (15), copy-number variants (16), gene gain (17, 18), or gene loss (19), targeting altered RNA expression of key AMR loci (20-23) provides a rapid way to identify the phenotypic consequences of the myriad genotypic variants that underpin AMR. For example, a quantitative real-time PCR (qPCR) targeting upregulation of three key AMR resistance-nodulation-division (RND) efflux pumps in the melioidosis pathogen, *Burkholderia pseudomallei*, enables the simultaneous identification of meropenem (MEM) resistance, and decreased susceptibility towards doxycycline and co-trimoxazole, in a single reaction (24).

Due to the large number of genetic mutations that can confer AMR in *P. aeruginosa* (25, 26), it remains impractical to target each individual variant using PCR. Fortunately, most clinically-relevant AMR in *P. aeruginosa* is thought to be conferred by a handful of key mechanisms, some of which alter gene expression to drive the AMR phenotype (25). The primary AMR-conferring mutations in *P. aeruginosa* include those that cause: i) *oprD* inactivation or repression, which commonly leads to carbapenem resistance (27, 28); ii) *ampC* hyper-production, a predominant cause of β-lactam AMR (25); iii) DNA gyrase A (*gyrA*) alteration, associated with fluoroquinolone (FQ) AMR (13, 29); and iv) upregulation of one or more RND efflux pumps, leading to AMR or MDR (23). The most important AMR-conferring RND efflux pumps in *P. aeruginosa* are MexAB-OprM (30) (FQs, β-lactams including carbapenems (31), and β-lactam inhibitors (25)), MexEF-OprN (FQs) (32), MexXY (33) (aminoglycosides and FQs (34)), and MexCD-OprJ (35) (FQs (36) and some fourth-generation cephalosporins (37)).

Whilst PCR assays exist to individually detect differential gene expression of *ampC* (20, 26, 34), *mexB* (22, 38), *mexX* (20, 39), *mexY, mexD, mexF* (22, 23), *mexA* and *oprD* (22, 34, 39) in *P. aeruginosa*, none simultaneously detect expression of these genes. In addition, nearly all *P. aeruginosa* AMR gene expression studies to date have assessed lab-cultured isolates (23, 38); with only one published study assessing AMR gene expression *in vivo*, whereby singleplex assays were used to detect *ampC* and *mexX* expression in 31 CF sputa, followed by comparison with corresponding derived isolates (20).

To address these knowledge gaps, we developed and validated two multiplex qPCR assays to simultaneously detect altered expression of six key genes that confer AMR towards most clinically-relevant antibiotic classes, using *rpsL* as an internal reference gene for expression normalisation. qPCRs were initially tested in cultures, and subsequently directly on 15 sputa, to permit *in vivo P. aeruginosa* gene expression characterisation (20), thus removing transcriptional biases introduced by culturing. qPCR results were also correlated with current anti-pseudomonal treatment to determine the impact of antibiotic therapy on *in vivo* AMR gene expression, along with strain phenotype and predicted *in silico* AMR genotype data.

## Methods

### Ethics statement and participant recruitment

This study was approved by The Prince Charles Hospital (TPCH) Human Research Ethics Committee, project IDs HREC/13/QPCH/127 (CF) and HREC/2019/QPCH/48013 (COPD). Site-specific approvals were obtained for CF recruitment at TPCH, Brisbane, Australia, and COPD recruitment at Sunshine Coast Hospital and Health Service, Sunshine Coast, Australia. CF sputa were collected from participants with chronic *P. aeruginosa* infection who were admitted to the Cystic Fibrosis Centre at TPCH for intravenous antibiotic therapy; COPD sputa was collected from participants either admitted to Sunshine Coast University Hospital (SCUH) for pulmonary exacerbations, or whose symptoms were being managed at home as part of the Respiratory Acute Discharge Service program; whether they had acute or chronic *P. aeruginosa* infection was not known. All participants provided written consent.

### Total RNA extraction and microbial RNA enrichment from sputa

Nine sputa from five CF participants, and six sputa from six COPD participants (40) (Table 1), all with high-load *P. aeruginosa* infections according to sputum culture onto MacConkey agar (Oxoid, Heidelberg West, VIC, Australia), were subjected to total RNA extraction to determine *in vivo P. aeruginosa* AMR gene expression profiles. An additional six sputa from *P. aeruginosa*-infected COPD participants with low *P. aeruginosa* load were also examined, along with three COPD sputa and 1 COPD bronchial washing from *P. aeruginosa* culture-negative participants. Sputa were collected directly into RNA stabilisation reagent (10mM EDTA, 25mM sodium citrate, 700g/L ammonium sulphate; pH=5.2; Sigma-Aldrich, Castle Hill, NSW, Australia) to immediately halt transcriptional activity, and kept at 4□C until RNA extraction (up to ∼1 month). Between 30 μL and 1 mL sputum was placed into 0.5-6 mL TRI Reagent LS (Sigma) and 30-240 μL 2-mercaptoethanol (Sigma) aliquoted into sterile, RNase-free 2 mL O-ring tubes (SSIBio, Lodi, CA, USA) containing ∼100 μL equal mixture of 0.1 and 0.5 mm zirconia beads (Daintree Scientific, St Helens, Tasmania, Australia). Tubes were parafilmed and subjected to four rounds of bead beating at 30 sec pulses using the Minilys tissue homogeniser on the medium setting (Bertin Instruments, Montigny-le-Bretonneux, France), with samples cooled on ice for >30 sec after each round to minimise RNA degradation. Extracted RNA was treated with DNase Max (Qiagen), confirmed DNA-free using a 16S ribosomal RNA gene PCR (41), and host RNA-depleted using MICROBEnrich (Thermo Fisher Scientific, Seventeen Mile Rocks, QLD, Australia) as per manufacturer’s instructions. cDNA (iScript; Bio-Rad, Gladesville, NSW, Australia) was qPCR-tested at neat and 1:10 concentrations.

**Table 1.**
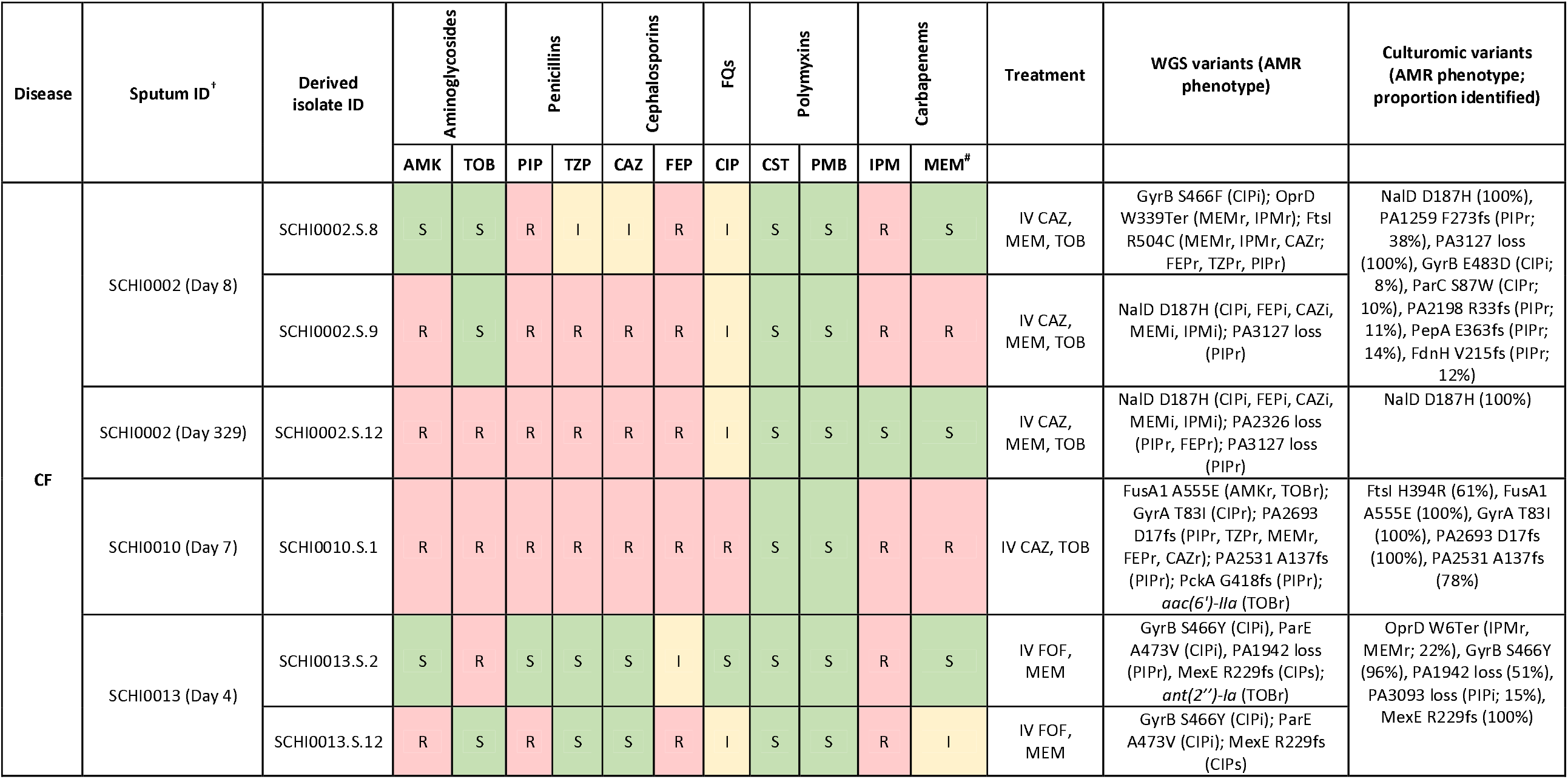
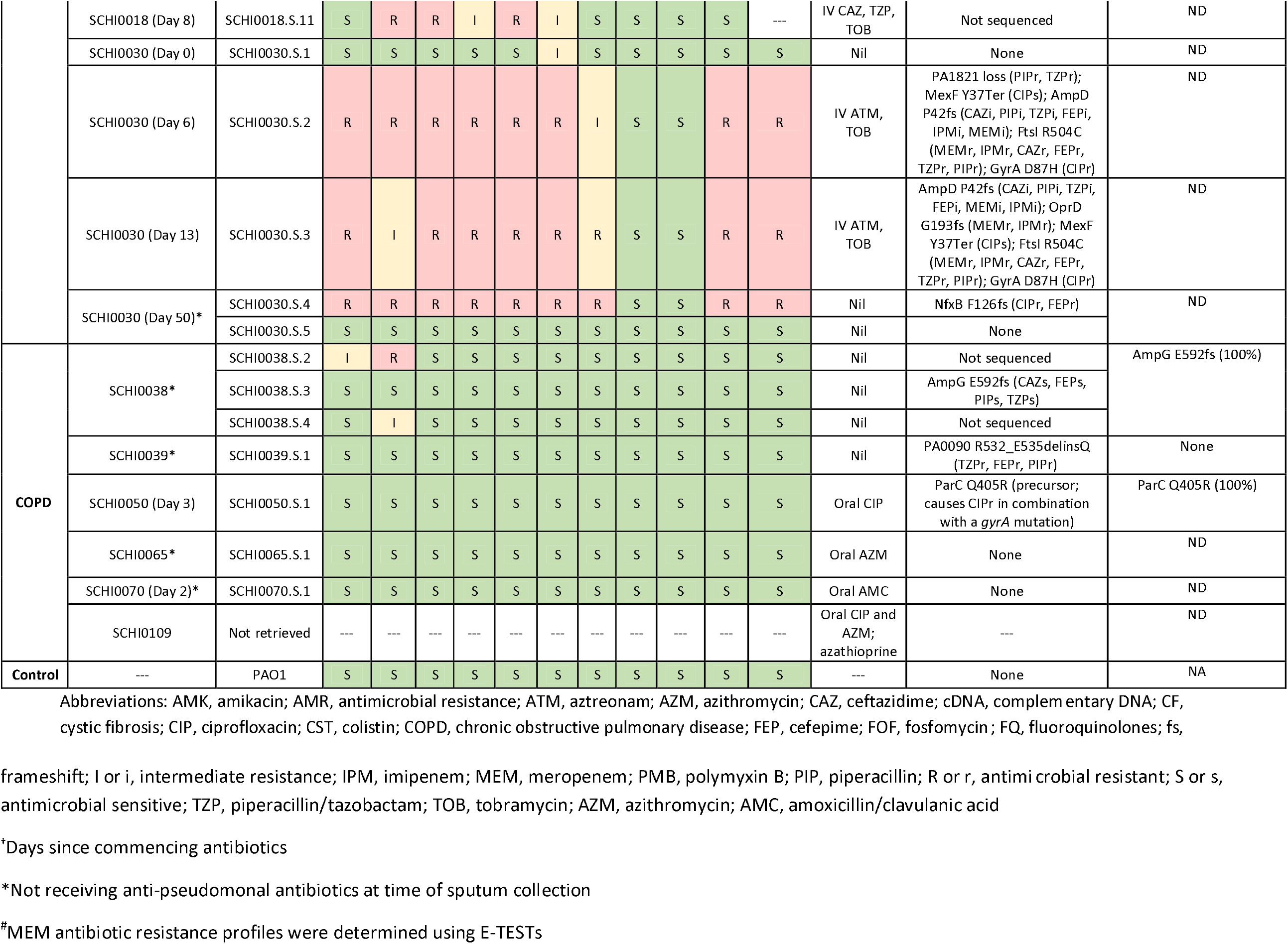
*Pseudomonas aeruginosa*-positive sputa, corresponding derived isolates, disc diffusion profiles across 11 anti-pseudomonal antibiotics, and predicted antimicrobial resistance (AMR) phenotypes from whole-genome sequencing (WGS) and culturomic data.

### Growth conditions and antibiotic sensitivity testing

Nineteen *P. aeruginosa* strains (12 CF, 7 COPD) were isolated from sputa using MacConkey agar and incubated at 37□C for 24 h. Subcultures were grown on Luria-Bertani (LB) agar (Oxoid) under the same conditions. All strains were confirmed as *P. aeruginosa* by rapid chelex-100 (Bio-Rad) heat soak extraction (42) followed by *ecfX* real-time PCR on a 1:50 dilution of the chelex supernatant (43). Susceptibility towards amikacin (AMK; 30μg), cefepime (30μg), ceftazidime (CAZ; 30μg), ciprofloxacin (5μg), colistin (10μg), imipenem (10μg), piperacillin (30μg), piperacillin-tazobactam (100μg/10μg), polymyxin B (300U), and tobramycin (TOB; 10μg) were determined by disc diffusion (Edwards Group, Murrarie, QLD, Australia); Etests were used for MEM susceptibility testing (bioMérieux, Baulkham Hills, NSW, Australia). Sensitivity, intermediate resistance, and AMR were determined using CLSI M100S-Ed27:2021 guidelines (Table 1). PAO1 (LMG 12228) was included as a wild-type reference. MDR was defined as non-susceptibility to at least one antibiotic in three or more antibiotic classes (44).

### Growth experiments and RNA extraction

To assess AMR gene expression levels in derived cultures, a starting inoculum of 10^5^ cells was grown to late-log phase (OD_600_ ≈1) with orbital shaking at 250 rpm (Ratek Laboratory Equipment [model number OM11], Boronia, VIC, Australia) for 16 h at 37□C in 2mL LB broth ±2.5% NaCl. RNA was extracted using TRI Reagent (Sigma-Aldrich) and treated with TURBO DNase (Thermo Fisher Scientific) according to manufacturer’s instructions. 1:10-diluted DNA-free RNA was converted to cDNA using iScript. cDNA was diluted 1:25 in molecular-grade H_2_O prior to qPCR.

We also attempted to induce AMR gene expression in two *P. aeruginosa* cultures, SCHI0002.S.9 and SCHI0010.S.1, using sub-inhibitory concentrations (0.25µg/mL each) of five antibiotics not used in *P. aeruginosa* treatment but known to induce its RND efflux pumps – gentamicin for *mexX* upregulation (45), novobiocin for *mexB* upregulation (31), norfloxacin for *mexC* upregulation (46), chloramphenicol for *mexE* upregulation (47) and ampicillin for *ampC* upregulation (48). These strains were selected as both encode MDR phenotypes; in addition, SCHI0010.S.1 encodes a high-consequence frameshift mutation in MutS (R360fs), resulting in a hypermutator phenotype (49).

### Multiplex AMR locus qPCR assay design and PCR conditions

To quantify *mexAB-oprM, mexCD-oprJ, mexEF-OprN, mexXY-OprM, ampC*, and *oprD* expression, two multiplex assays targeting the *mexB, mexC, mexE, mexY, ampC*, and *oprD* genes, respectively, were designed (Table 2). A BLAST database comprising 730 *P. aeruginosa* genomes (50-54) was used to identify conserved regions for oligo design. Oligo self-dimers and heterodimers were assessed and avoided as previously described (55). Each PCR consisted of 1× SsoAdvanced Universal Probes Supermix (Bio-Rad), optimised primer and probe concentrations (Macrogen Inc., Geumcheon-gu, Seoul, South Korea; Table 2), 1 µL genomic DNA (gDNA) or cDNA template, and PCR-grade H_2_O, to 5 µL. Isolate gDNA/cDNA thermocycling comprised enzyme activation at 95□C for 2 min, followed by a 2-step program (95□C for 5 sec and 60□C for 15 sec) for 45 cycles; for sputum cDNA, a 3-step program (95□C for 5 sec, 60□C for 15 sec, and 72□C for 15 sec) for 45 cycles was used to enhance *P. aeruginosa* detection from polymicrobial DNA.

**Table 2.**
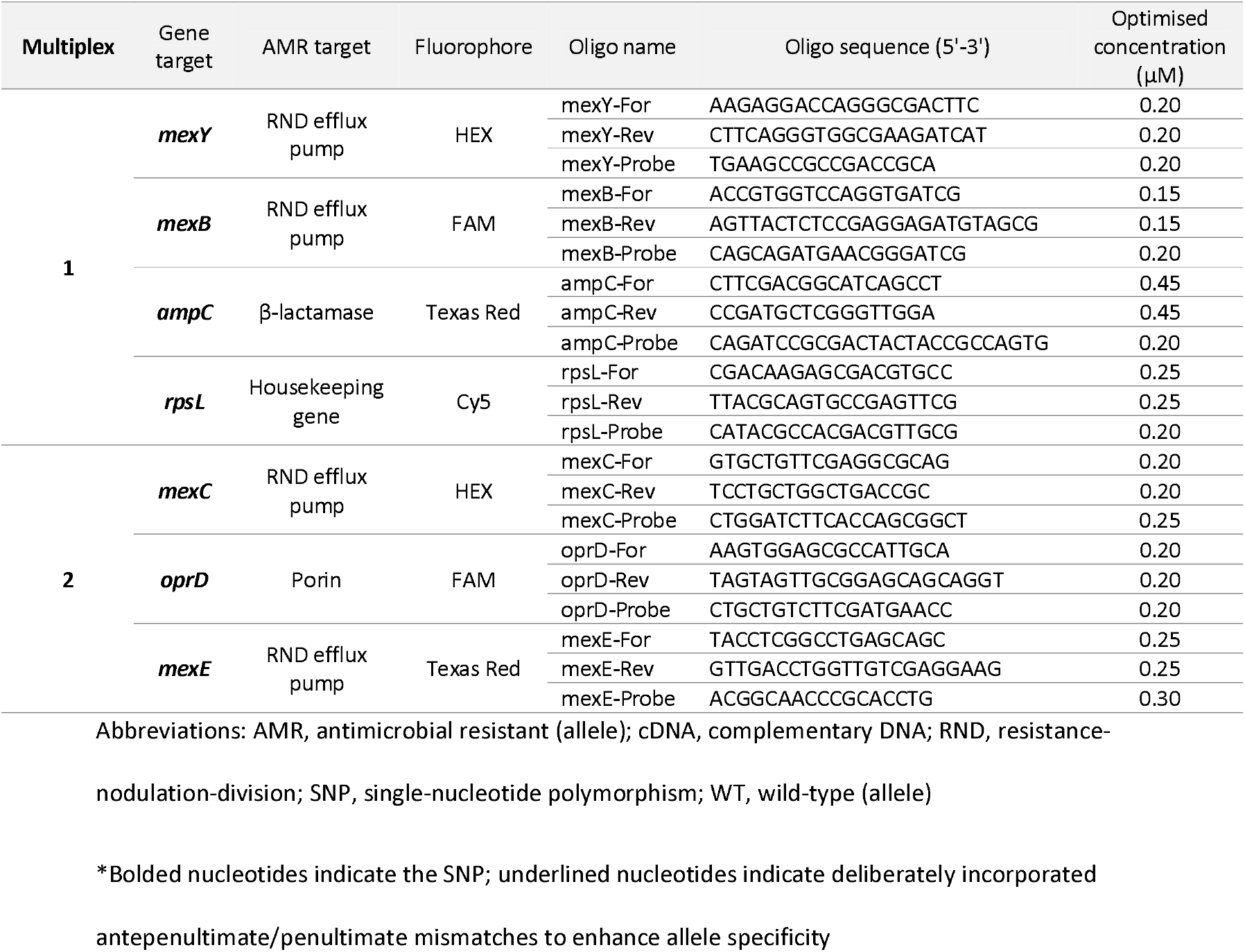
Primers and probes designed in this study.

### qPCR assay limits of detection (LoD) and quantification (LoQ)

LoD and LoQ values were determined for each assay when in multiplex format using 1:10 serial dilutions of SCHI0005.S.10 *P. aeruginosa* genomic DNA ranging from 40 to 4 × 10^−6^ ng across eight replicates, as previously described (56, 57). Genome equivalents (GEs) were calculated based on the average PAO1 genome size of 6.22Mbp and a single copy of each gene. For all assays, all samples were run in duplicate, and at least two no-template controls were included in each run.

### qPCR normalisation and analysis

The constitutively expressed *rpsL* gene (Table S1) was used to normalise AMR gene expression via the difference in cycles-to-threshold (ΔC_T_) method (34, 58). After *rpsL* normalisation, ΔC_T_ values were further normalised against either PAO1 expression, or to SCHI0070_D2 COPD sputum (ΔΔC_T_), the latter of which enabled gene expression comparison among sputa, and between sputa and derived isolates. SCHI0070_D2 COPD sputum was chosen for normalisation as this participant had no recent history of anti-pseudomonal antibiotic treatment, its derived isolate, SCHI0070.S.1, was sensitive towards all 11 anti-pseudomonal antibiotics (Table 1), and there was good gene expression across all seven loci for this sample, thus enabling relative expression profiling of qPCR-tested genes across all tested sputa. For AMR loci lacking detectable PCR amplification, a C_T_ value of 45 was assigned to permit visualisation of genes with very low/no expression. According to previously defined criteria (23), strains were regarded as hyper-producers if their AMR loci exhibited >10-fold higher expression compared with PAO1, except for *mexB* and *mexY*, which were considered hyper-producing if >3-fold higher expression than PAO1 (23, 34, 38).

### Strain genome sequencing

The genomes of 14 strains have been previously published (13); paired-end Illumina genomes for SCHI0065.S.1 and SCHI0070.S.1 were generated in the current study and appended to NCBI BioProject PRJNA761496. We did not sequence SCHI0018.S.11, SCHI0038.S.2, or SCHI0038.S.4 as these data were not considered essential for study outcomes.

### Culturome sequencing and *P. aeruginosa* mixture analysis

Culturomes for SCHI0002_D8, SCHI0002_D329, SCHI0010_D7, SCHI0013_D4, SCHI0038, SCHI0039, and SCHI0050_D3 were generated from Chocolate agar culture sweeps. Microbial cells were lysed with 250 U/mL mutanolysin, 22 U/mL lysostaphin, and 20mg/mL lysozyme, followed by DNA extraction using the Qiagen DNeasy Blood and Tissue kit ‘Pretreatment for Gram-Positive Bacteria’ protocol. Culturomic data were Illumina paired-end sequenced at the Australian Genome Research Facility (Melbourne, VIC, Australia) or Ramaciotti Centre for Genomics (Sydney, NSW, Australia) to obtain a minimum of 7 million reads. The culturomic sequence data are available under NCBI BioProject PRJNA761496. *P. aeruginosa* mixtures were assessed by mapping reads to PAO1 (GenBank: CP053028.1) with ARDaP v1.9 (59) (https://github.com/dsarov/ARDaP) using the --mixtures flag. Heterogeneous variant sites were then identified and counted from each sample’s variant call file (.vcf) based on ‘0/1’ in the ‘GT’ (genotype) field.

### AMR prediction from genomes and culturomes

*In silico* AMR prediction for 16 genome-sequenced isolates, PAO1 (Table 1), and the sputum culturomic data was carried out using the *P. aeruginosa-* specific database in ARDaP. Prior to ARDaP analysis, *Pseudomonas* (taxid: 286) and *P. aeruginosa* (taxid: 287) specific reads were extracted from the raw culturomic data using Kraken 2 (60) to classify reads followed by sektq (https://github.com/lh3/seqtk) for retrieval, with resultant coverage ranging from 21x to 43x (calculated using Mosdepth (61)). For culturomic analysis, ARDaP was again run in mixture mode (--mixtures flag), with the ResFinder (62) component disabled due to the presence of multiple species.

### Statistical analysis

Gene expression differences were assessed using a two-way analysis of variance test and Tukey’s multiple comparison test using GraphPad Prism (GraphPad Software Inc., CA, USA). Corrected *p* values of <0.05 were considered significant.

## Results

### Multiplex qPCR assay performance

To assess assay sensitivity, LoD and LoQ values were determined using a 10-fold DNA dilution series (40 ng to 0.04 fg). The LoD for all assays was 400 fg (59 GEs), the LoQ for *mexB, mexE* and *mexC* was 400fg (59 GEs), and the LoQ for *rpsL, ampC, mexY* and *oprD* was 4000 fg (587 GEs) (Figures S1 and S2). Based on these results, an *rpsL* C_T_<30 was identified as the upper threshold for accurately assessing gene expression of AMR loci.

### Antibiotic susceptibility profiles

Testing towards 11 anti-pseudomonal antibiotics identified much higher rates of AMR (range: 0-9 antibiotics) and MDR (10/12 strains) in CF-derived isolates when compared with COPD-derived isolates (range: 0-2 antibiotics; 0/7 MDR strains) (Table 1). This was consistent with much greater antibiotic use in the CF cohort (Table 1) (40). The lowest susceptibility rates were towards cefepime (0% CF and 100% COPD) and piperacillin (33% CF and 100% COPD), whereas all 19 strains were susceptible to colistin and polymyxin B (Table 1). Interestingly, cefepime is rarely used for CF treatment at TPCH, indicating probable cross-resistance towards this antibiotic in all CF-derived isolates. None of the participants were receiving polymyxins; as such, no AMR was expected towards these antibiotics.

### In silico AMR prediction in isolate genomes

Genomic analysis of the derived *P. aeruginosa* isolates revealed a much greater number of AMR determinants in the CF-derived strains (Table 1). Although some AMR determinants were identified in COPD isolates, only one was associated with dysregulated expression of the six AMR loci. In contrast, many CF-derived isolates encoded mutations predicted to alter gene expression of our qPCR gene targets (Table 1). Specifically, *ampC* overexpression was predicted in SCHI0030.S.2 and SCHI0030.S.3 due to AmpD regulator loss (P42fs) (63), and a non-inducible *ampC* phenotype was predicted in SCHI0038.S.3 due to a E592fs nonsense mutation in AmpG (64); *mexAB-oprM* overexpression was predicted in SCHI0002.S.9 and SCHI0002.S.12 due to a D187H missense mutation in the NalD regulator (65); and *mexCD-oprJ* overexpression was predicted in SCHI0030.S.4 due to NfxB regulator loss (F126fs) (66). No mutations associated with *mexEF-oprN* or *mexXY-oprM* overexpression were identified. Although not associated with gene expression changes, *oprD* loss was predicted in SCHI0002.S.8 (W339Ter) and SCHI0030.S.3 (G193fs), which is linked to carbapenem resistance, especially when associated with *ampC* upregulation (25, 67) as observed in SCHI0030.S.3.

Other AMR determinants were identified in our strain dataset, including multiple determinants encoding decreased susceptibility or AMR towards CIP (GyrA T83I, D87H or D87N, GyrB S466F and ParE A473V (68)) (Table 1). In addition, an R504C missense variant in FtsI, known to confer resistance to β-lactams (69), was detected in three CF-derived, multi-β-lactam-resistant isolates (SCHI0002.S.8, SCHI0030.S.2, and SCHI0030.S.3), and a FusA1 missense mutation, A555E (70), was identified in CF-derived isolate SCHI0010.S.1 (AMK- and TOB-resistant) (Table 1). Finally, MexEF-OprN loss was predicted in SCHI0013.S.2 and SCHI0013.S.12 due to a frameshift (R229fs) in the MexE membrane fusion protein, and in SCHI0030.S.2 and SCHI0030.S.3 due to the introduction of a stop codon (Y37Ter) in the MexF multidrug inner membrane transporter (71). Unlike the AMR variants, these *mexE* and *mexF* mutations render the MexEF-OprN pump inoperable, and thus unable to contribute towards AMR, irrespective of *mexEF-oprN* upregulation.

### In silico AMR prediction in culturomes

Mixed variant site analysis (Table S2) of culturomic data identified multiple *P. aeruginosa* strains in 1/4 CF and 0/3 COPD sputa. The SCHI0002_D8 culturome had a high number of mixed variant sites (21664 variants), confirming that ≥2 genetically distinct strains were present; in contrast, multiple strains were not detected in the SCHI0002_D329, SCHI0010_D7, SCHI0013_D4, SCHI0038, SCHI0039, or SCHI0050_D3 sputa (833, 1521, 701, 1039, 688, 678 variants, respectively) when compared with mixed variant site analysis in clonal, derived strains (Table S2).

ARDaP AMR prediction (Table 1) identified AMR determinants not retrieved during single-strain culturing in 3/7 culturomes (3/4 CF and 0/3 COPD sputa). GyrB E483D (72) and ParC S87W (73), known to contribute to ciprofloxacin resistance, were identified in SCHI0002_D8 at a frequency of 8% and 10%, respectively; further, this sample had minor components of the AMR determinants FdnH V215fs (12%), PA1259 F273fs (38%), PA2198 R33fs (11%), and PepA E363fs (14%), all of which cause decreased piperacillin susceptibility (74). A FtsI H394R missense mutation (75) was identified in SCHI0010 at 61% prevalence, and in SCHI0013, a W6Ter nonsense mutation caused by a G→A transition was identified in OprD at 22% prevalence, predicted to cause carbapenem resistance, along with PA3093 loss (15%), associated with intermediate piperacillin resistance (Table 1). Importantly, no additional mutations were identified that were predicted to alter expression of the six AMR loci. Further, the culturomic data did not detect some AMR determinants found in the single-isolate data (Table 1).

### Comparison of AMR variant prediction with *in vitro* AMR gene expression

Relative AMR gene expression in derived *P. isolates* grown in LB broth (*n*=15; Figure S3), LB+NaCl (*n*=14; Figure S4), or a 5-antibiotic cocktail (*n*=2; Figure S5) was assessed. All qPCR targets exhibited expression in at least one strain; however, in some strains, particularly the CF-derived isolates, certain genes exhibited no expression (Figures S3-S5). Just two loci were expressed in all tested strains across all conditions: the housekeeping gene *rpsL* (C_T_=25.3±1.97) and *mexB* (C_T_=29.0±2.14). *oprD* was expressed in all strains except SCHI0039.S.1 grown in LB+NaCl. In contrast, *ampC*, and *mexC, mexE*, and *mexY* were expressed in 1, 23, 26, and 2 of the 31 tested strains, respectively. Genetic analysis of all qPCR-targeted loci confirmed 100% primer and probe matches in those strains with genomic data, thereby ruling out assay failure as a cause of undetectable expression.

Unexpectedly, there was no concordance between *in silico* AMR predictions and qPCR results for strains grown under any of the tested conditions. SCHI0030.S.2 and SCHI0030.S.3, which were predicted to upregulate *ampC* through a previously undocumented AmpD frameshift variant (P42fs), exhibited negligible or no *ampC* expression when grown in LB broth or LB+NaCl (Figures S3 and S4). In contrast, SCHI0010.S.1, which does not encode any known chromosomal variant associated with *ampC* upregulation (Table 1), demonstrated *ampC* hyper-expression (43x) in LB broth relative to PAO1, although no detectable *ampC* expression under salt (Figure S2) or antibiotic stress (Figure S3). Similarly, SCHI0002.S.9 and SCHI0002.S.12 failed to overexpress *mexB*, and, surprisingly, SCHI0030.S.4 failed to overexpress *mexC*, despite predicted upregulation. Once again, SCHI0010.S.1 grown in LB, but not in LB+NaCl, unexpectedly hyper-expressed *mexB* (19x), *mexC* (19x), *mexE* (24x), and *mexY* (16x) in lieu of encoding any known variants associated with upregulation of these loci. No other strain tested under any condition upregulated *mexB, mexC, mexE*, or *mexY*. Although no strains were predicted to exhibit *oprD* downregulation, *oprD* was not expressed in SCHI0039.S.1 grown in LB±NaCl, and was significantly downregulated in SCHI0010.S.1 grown in LB+NaCl (Figures S3 and S4). All other strains exhibited good *oprD* expression, including SCHI0002.S.8 and SCHI0030.S.3; although these strains encode truncated OprD, these deleterious mutations occur downstream of the oligo-binding sites, and are therefore undetected by our assay.

### AMR gene expression in sputa

All COPD sputa yielded more similar expression profiles to the SCHI0070_D2 COPD control than any of the nine CF sputa (Figure 1). The only overexpressed loci in the CF or COPD sputa were: *ampC* (4/9 CF [1.7-15.1x] and 2/5 COPD [1.7-3.5x]), *mexB* (3/5 COPD [1.5-2.5x]), *mexC* (2/9 CF [1.7-2.1x] and 2/5 COPD [6.4x each]), *mexY* (4/5 COPD [1.8-7.3x]), and *oprD* (3/5 COPD [1.7-51x]) (Figure 2). All CF sputa exhibited downregulation or no expression of *mexB* (≥4.0x), *mexE* (≥3.9x), *mexY* (≥5.2x), and *oprD* (≥377x), and 6/9 were downregulated or had no detectable expression for *mexC* (≥3.4x). When comparing CF and COPD sputum expression, no significant difference was observed for *ampC, mexB*, or *mexC*; in contrast, *mexE, mexY*, and *oprD* were all significantly downregulated in CF sputa (Figure 1).

**Figure 1.**
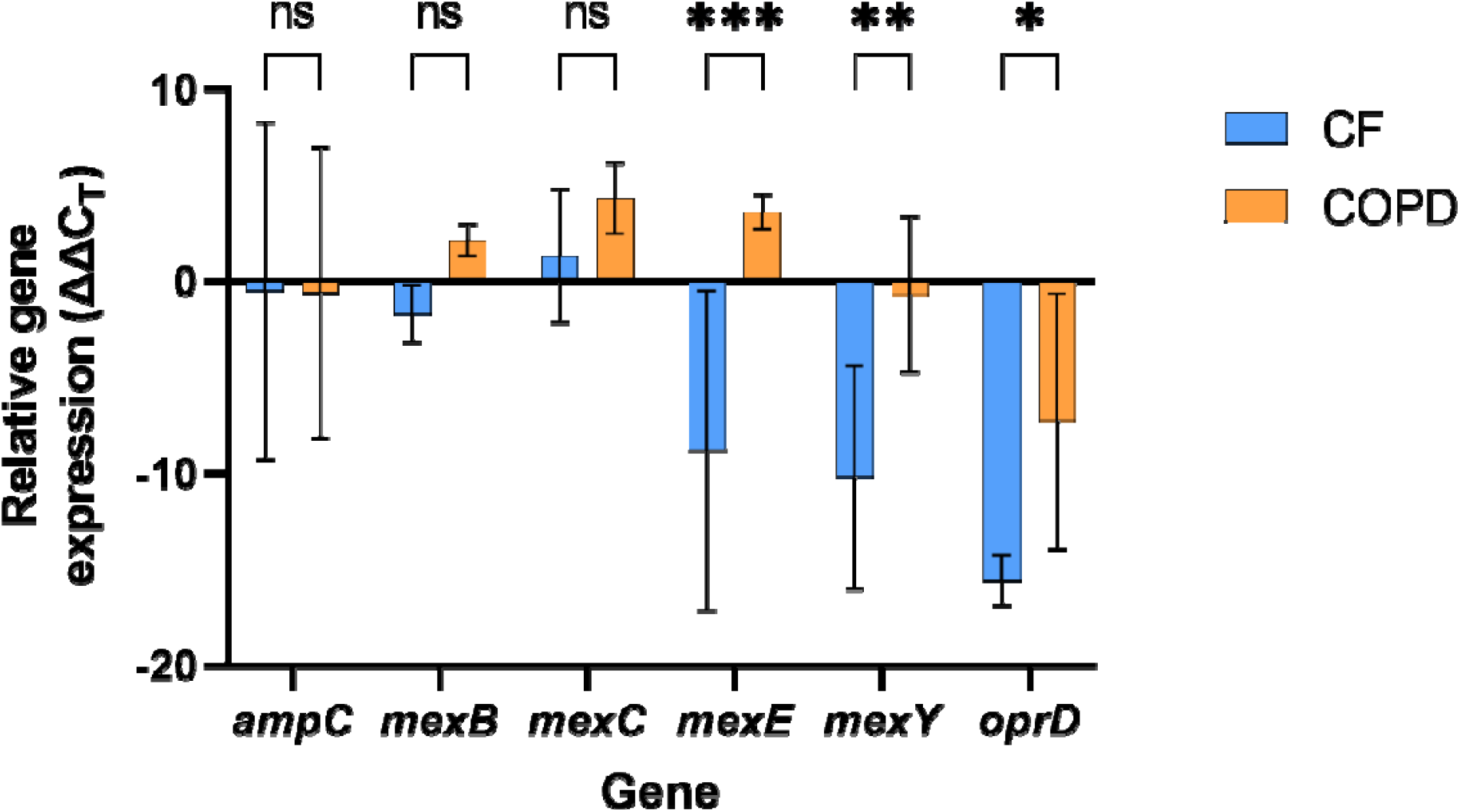
*In vivo* expression of *Pseudomonas aeruginosa* antimicrobial resistance loci in six cystic fibrosis (CF) and five chronic obstructive pulmonary disease (COPD) sputa. Gene expression for each sample at each locus was first normalised against *rpsL* (ΔC_T_); relative log_2_ fold-change expression was then determined by comparing ΔC_T_ values against COPD participant SCHI0070 (i.e. ΔΔC_T_), who was not receiving anti-pseudomonal antibiotics at the time of sputum collection. For SCHI0030, in which longitudinal sputa over antibiotic treatment were obtained, only the Day 0 sputum was used in this analysis to ensure independence of all observations. *, adjusted *p* <0.05; **, adjusted *p* <0.01; ***, adjusted *p* <0.001; ns, not significant. Error bars=95% confidence interval.

**Figure 2.**
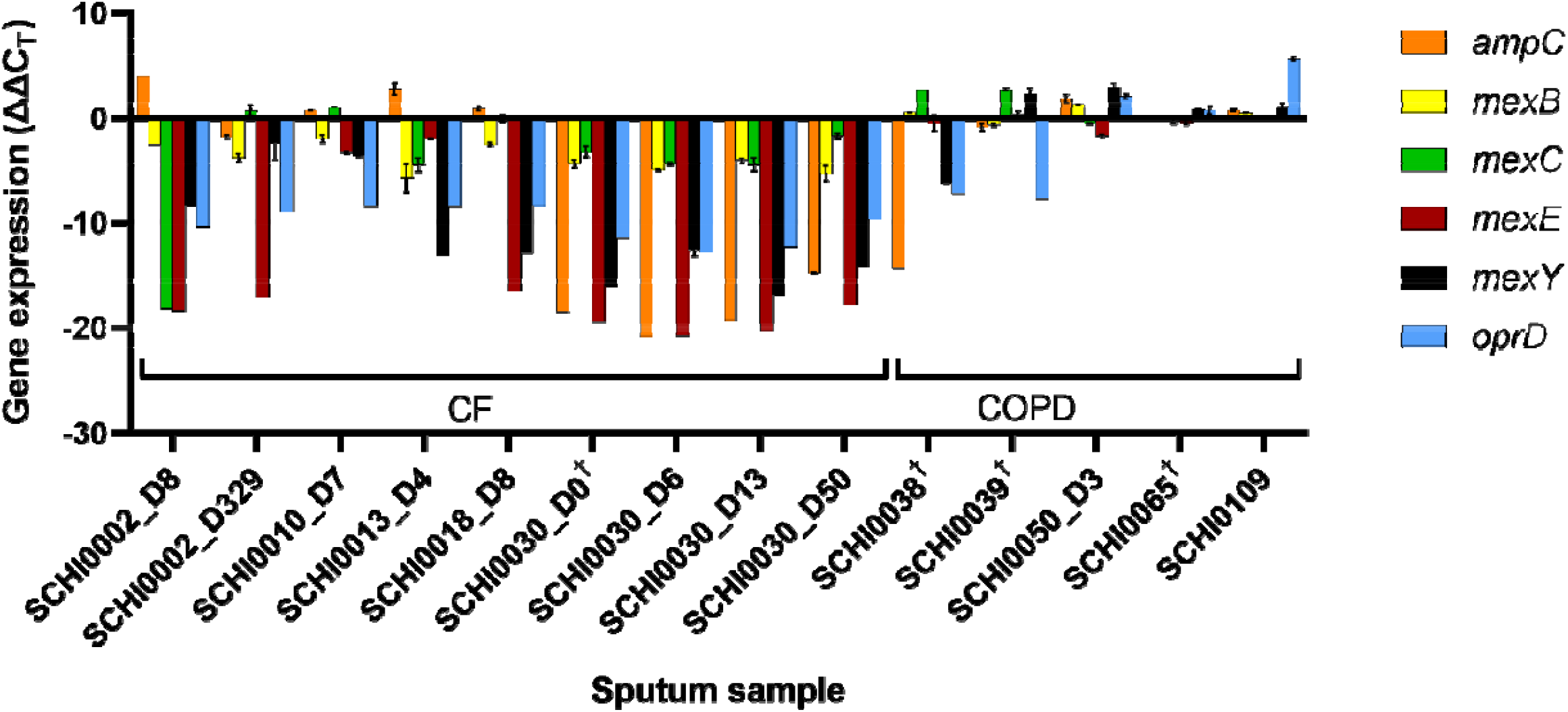
Differential expression (relative log_2_ fold change) of *Pseudomonas aeruginosa* antimicrobial resistance loci in cystic fibrosis (CF) and chronic obstructive pulmonary disease (COPD) sputa. Gene expression for each locus was first normalised against *rpsL* (ΔC_T_); relative expression was then determined by comparing ΔC_T_ values against sputum from COPD participant SCHI0070 (i.e. ΔΔC_T_), who was not receiving anti-pseudomonal antibiotics at the time of sputum collection. For participants receiving anti-pseudomonal antibiotics, sputa are denoted by ‘Day post-antibiotics’ (D) (see Table 1 for details). Error bars, standard deviation. †Participants receiving non-pseudomonal antibiotics at the time of sputum collection.

Correlation of qPCR results with antibiotic treatment (Table 1) identified that sputum *ampC* expression levels were a good indicator of current β-lactam use, with 4/5 sputa with elevated *ampC* expression collected from participants contemporaneously receiving β-lactams (SCHI0002_D8, SCHI0010_D7, SCHI0013_D4, and SCHI0018_D8). Notably, 2/3 sputa (SCHI0002_D8 and SCHI0013_D4) obtained from participants being treated with MEM, either with or without CAZ, overexpressed *ampC* at high levels (6.7-15x); in contrast, *ampC* expression was only moderately upregulated (1.7-1.9x) in two sputa (SCHI0010_D7 and SCHI0018_D8) from participants on CAZ but not MEM (Table 1; Figure 2). *ampC* was downregulated or not expressed in eight sputa (Figure 2) - of these, four were collected from participants receiving no antibiotics at the time of collection, three were receiving antibiotics other than β-lactams, and just one (SCHI0002_D329) was receiving CAZ and MEM at the time of sputum collection (Table 1).

### AMR gene expression between sputa and *in vitro* cultures

Sputum AMR gene expression was compared with derived cultures grown in LB broth (Figure S3), LB+NaCl (Figure S4), and a 5-antibiotic cocktail (Figure S5) to determine whether culture AMR gene expression was reflective of *in vivo* conditions. Overall, there was very poor concordance between *in vivo* and *in vitro* expression profiles across all tested *in vitro* conditions, with most loci exhibiting dissimilar expression between derived isolates and their corresponding sputa. Most striking was *oprD*, which, in most cases, was upregulated in strains grown under the three *in vitro* conditions (Figures S3-S5) yet largely downregulated *in vivo* (Figure 2).

### RND efflux pump expression during antibiotic treatment

The four longitudinal sputa collected from SCHI0030, who received intravenous TOB and aztreonam for 15 days, provides an interesting case study for observing AMR gene expression dynamics before, during, and after antibiotic treatment (Table 1). Although TOB has previously been shown to induce MexXY-OprM expression in CF sputa (20), we unexpectedly did not observe AMR gene expression changes in SCHI0030 sputa (Figure 2). Five sputa from other CF participants also showed limited or no *mexY* expression. In comparison, *mexY* was significantly upregulated in 4/5 COPD sputa (Figure 2), none of whom have ever received TOB (Table 1).

## Discussion

Here, we developed and validated two multiplex qPCR assays for the simultaneous detection of altered AMR locus expression in the ESKAPE pathogen, *P. aeruginosa*. When run in multiplex format, our qPCRs showed very good to excellent LoD and LoQ values. qPCRs can be used both on *P. aeruginosa* cultures and directly on human RNA-depleted sputa, the latter of which provides *in vivo* expression profiling that overcomes transcriptional bias introduced with *in vitro* culture (20). Building upon the pioneering work of Martin and colleagues, who first reported limited correlation in *ampC* and *mexX* expression between sputa and corresponding derived isolates (20), we also observed poor concordance between *in vivo* and *in vitro* expression profiles across six tested AMR loci. Further, we found surprisingly poor correlation between *in silico*-predicted and *in vitro* AMR gene expression. These findings raise questions about the clinical utility of *in vitro* antimicrobial gene expression profiling and highlight the need for more work in this critical area of AMR research.

### AmpC-driven AMR

We found that sputum *ampC* expression correlated well with contemporaneous β-lactam treatment. In particular, two CF participants (SCHI0010 and SCHI0018) receiving CAZ but no other β-lactams overexpressed *ampC* at moderate levels (1.7-2x); this overexpression increased to 6.7-15x in two other participants (SCHI0002 and SCHI0013) receiving MEM (Table 1; Figure 2). Carbapenems such as MEM can be potent *ampC* inducers (67) and can remain effective even when *ampC* expression is elevated (25), whereas cephalosporins such as CAZ are relatively weak inducers of *ampC* and are readily hydrolysed by this enzyme (25). Our findings confirm that MEM can act as a strong inducer of *ampC* expression *in vivo*. In light of this, we recommend that *P. aeruginosa* infections should not simultaneously be treated with carbapenems and cephalosporins due to potential *ampC* induction, which would render cephalosporin (and likely other β-lactam) antibiotics ineffective (25). In contrast, eight sputa exhibited downregulated or no *ampC* expression (Figure 2), seven of which were taken from participants either not on antibiotics or not receiving β-lactams at the time of sample collection, which likely explains the lack of β-lactam-driven *ampC* selection pressure. Of note, two COPD sputa, SCHI0109 and SCHI0050_D3, showed *ampC* upregulation (1.7-3.5x), despite neither participant receiving contemporaneous β-lactam therapy. Whilst the basis for this upregulation is unknown, we speculate that β-lactam use in these participants would be ineffective for *P. aeruginosa* eradication. Taken together, our results demonstrate that routine *ampC* qPCR testing of *P. aeruginosa-*positive specimens is a useful diagnostic for monitoring *in vivo* cephalosporin efficacy, enabling more rapid treatment shifts in instances where *ampC* is upregulated.

Unlike sputa, almost all derived isolates failed to express *ampC* when grown in three different liquid media. The exception was the derived isolate from CF participant SCHI0010. In LB broth, SCHI0010.S.1 exhibited *ampC* hyper-expression (15x relative to SCHI0070_D2, and 43x relative to PAO1; Figure S3), yet the corresponding sputum showed only modest *ampC* upregulation (1.7x; Figure 2). This finding was unexpected as SCHI0010.S.1 does not encode any known chromosomal variant associated with *ampC* upregulation (Table 1), and did not overexpress *ampC* when grown in LB+NaCl or the 5-antibiotic cocktail (Figures S4-S5). These results indicate that SCHI0010.S.1 encodes an enigmatic determinant for upregulating *ampC* expression when grown under particular conditions. Whilst the basis of this overexpression requires further investigation, we postulate that it may be driven by global transcriptional regulator inactivation due to its hypermutator status, resulting in marked transcriptome differences affecting multiple gene pathways. In contrast, strains SCHI0030.S.2 and SCHI0030.S.3, both predicted to upregulate *ampC* due to a truncated AmpD, exhibited negligible or no *ampC* expression *in vitro* (Figures S3-S4). This result correlated with the longitudinal SCHI0030 sputa, all of which exhibited significantly downregulated *ampC* expression. One possibility for the lack of *ampC* expression in SCHI0030 is that the AmpD P42fs mutation may only confer a partially de-repressed *ampC* phenotype (26); another possibility is that additional chromosomal determinants, such as those occurring in other AmpD homologues (26) or DacB (76), may be required to confer *ampC* upregulation (63) in AmpD P42fs-encoding strains.

Expression of *ampC* was also undetectable in the MDR strain, SCHI0002.S.9, across the three *in vitro* conditions (Figures S3-S4). Whilst this observation is consistent with the lack of predicted *ampC* dysregulation in the SCHI0002.S.9 genome, it does not reflect the AMR phenotype of this strain, which was resistant to all six tested penicillin, cephalosporin, and carbapenem antibiotics (Table 1). In addition, the corresponding SCHI0002_D8 sputum displayed the greatest *ampC* upregulation (15x; Figure 2), probably due to contemporaneous CAZ and MEM treatment (Table 1). One possible explanation for these spurious results is that high sputum *ampC* levels are conferred by an un-retrieved *P. aeruginosa* strain (20), as supported by two distinct AMR phenotypes and genotypes isolated from SCHI0002_D8, SCHI0013_D4, and SCHI0030_D50 sputa (Table 1) and *in silico* strain mixture analysis of culturomic data, which identified multiple *P. aeruginosa* strains in one CF sputum sample (SCHI0002_D8) (Table S2). Indeed, phenotypic and genotypic diversity of *P. aeruginosa* is well-documented in chronic CF infections (77), and increasingly, in COPD infections (78). However, our culturomic mixture analysis failed to identify any additional mutations associated with altered gene expression of *ampC* or the other tested AMR loci (Table 1), so it remains unconfirmed whether undetected determinants are contributing to AMR. Another explanation is that *ampC* expression in SCHI0002 strains may require MEM (20) or cefoxitin (79) rather than ampicillin (48) for *in vitro* induction, as certain antibiotics are known to be better than others for inducing *ampC* (20). However, antibiotic induction only provides a crude representation of *in vivo P. aeruginosa* activity, as the lung environment is exceptionally difficult to mimic *in vitro* (20, 77, 80). The lack of correlation between *ampC* upregulation in sputa and derived cultures (Figures S3-S5) has also been reported in a separate study (20), and emphasises the challenges in replicating *in vivo* conditions *in vitro*.

### Efflux-mediated AMR

*mexB, mexC, mexE* and *mexY* represent the RND efflux pumps most frequently associated with AMR and MDR in *P. aeruginosa* (23, 25). As most CF participants were being treated with anti-pseudomonal antibiotics at the time of sputum collection, and many of their derived isolates encode key mutations associated with altered RND efflux pump expression (Table 1), we expected upregulation of these four efflux pumps in CF sputa. It was therefore surprising that *mexC* was the only efflux system that demonstrated modest (1.7-2.1x) upregulation in a few patients (2/9 CF sputa), whereas the other RND efflux systems were largely downregulated, or switched off entirely, in CF sputa (Figure 2). Also surprising was that most COPD sputa showed good expression of these loci, despite only two COPD participants contemporaneously receiving anti-pseudomonal antibiotics, and almost no AMR phenotypes or efflux pump expression-altering genotypes in the derived COPD isolates (Table 1). This difference between diseases was most apparent for *mexE, mexY*, and *oprD*, which were significantly downregulated in CF but not COPD sputa (Figure 1). Although our dataset is small, these observations suggest that RND efflux pump expression, and particularly *mexE* and *mexY* expression, may not be required for chronic *P. aeruginosa* persistence in CF airways. This finding contradicts prior *in vitro* work suggesting that MexXY-OprM efflux pump upregulation is necessary for *P. aeruginosa* survival in the CF lung (81). We further postulate that the MexEF-OprN and MexXY-OprM RND efflux pumps may have a minimal role, if any, in conferring AMR in *P. aeruginosa in vivo. mexE* and *mexY* qPCR testing across a larger CF and COPD sputum panel is needed to determine to veracity of our findings.

One possible explanation for efflux pump downregulation in CF sputa is that the CF cohort all harboured chronic *P. aeruginosa* infections, whereas the COPD cohort potentially only had transient (i.e. acute) infections. Like virulence factors (82), RND efflux pump expression may be upregulated during the acute phase but then downregulated once *P. aeruginosa* transitions to a chronic lifestyle, the latter of which is characterised by biofilm formation and reduced metabolic activity (25, 83). In support of this hypothesis, longitudinal sputa collected from CF participant SCHI0030 just prior, during, and after a 15-day course of intravenous TOB and aztreonam treatment showed virtually no change in RND efflux pump expression, with all AMR genes (including *mexY*) downregulated (Figure 2) despite the aminoglycoside onslaught. In addition, one of two SCHI0030 strains obtained 35 days after antibiotic cessation was fully susceptible to all 11 tested antibiotics, including TOB, hinting at insufficient antibiotic penetration due to biofilm. As ours is one of the few studies to investigate *P. aeruginosa* AMR gene expression *in vivo*, and the first to assess COPD sputa, more work is required to verify these hypotheses, as there remains stark limitations in our understanding of the characteristics of *P. aeruginosa* infection (including AMR development), particularly in people with COPD (84). Examining *in vivo* AMR gene expression dynamics over a long-term *P. aeruginosa* infection would shed important information about the adaptive processes that occur during the transition from acute infection to chronic persistence. We anticipate that our multiplex assays will be a valuable tool for such studies.

Analysis of RND efflux expression among the CF-derived isolates encoding predicted RND efflux upregulation revealed that almost none demonstrated increased efflux pump expression *in vitro* (Figures S3-S5). Curiously, the MDR and hypermutated isolate SCHI0010.S.1, which does not encode any known mutations associated with RND efflux pump upregulation, exhibited *mexB* (5.3x), *mexC* (1.7x), *mexE* (1.5x), and *mexY* (28x) overexpression when grown in LB, although not when grown in LB+NaCl or the 5-antibiotic cocktail (Figures S3-S5). These results indicate that there are growth-specific requirements associated with upregulation of these genes. Furthermore, these observations collectively indicate that *in vitro* efflux pump expression is not a good proxy for AMR phenotype, as our CF-derived *P. aeruginosa* isolates demonstrate extensive MDR, yet we were unable to reproduce their expected gene expression alterations *in vitro*. Our results contradict other *in vitro* experiments reporting RND efflux pump hyper-expression as a major contributing factor to AMR development in CF airway-derived *P. aeruginosa* (34, 39). Caution should thus be taken when attempting to correlate predicted AMR phenotypes with *in vitro* RND efflux pump expression profiles due to considerable AMR gene expression changes exhibited under different culture conditions.

### OprD-mediated AMR

As with the other AMR loci, we saw an unexpectedly poor correlation between *in vitro* and *in vivo oprD* expression. Whereas 17/19 *in vitro* cultures expressed *oprD* at high levels (Figures S3-S5), 9/9 CF and 2/5 COPD sputa exhibited no detectable *oprD* expression (Figure 2), and *oprD* was significantly downregulated in CF sputa (Figure 1). This finding strongly suggests that *oprD* downregulation is a common, and potentially dominant, adaptive *in vivo* trait in chronic *P. aeruginosa* infections. Another noteworthy observation is that *in vivo oprD* downregulation was not strictly associated with contemporaneous carbapenem use, as only three of the CF sputa were obtained during carbapenem treatment, and none of the COPD participants had ever received carbapenems. We posit that this adaptive mechanism may provide a rapid, efficient, and effective way for *P. aeruginosa* to evade carbapenems in lieu of encoding functional *oprD* loss. In support of this hypothesis, two prior studies reported decreased *oprD* expression as a cause of carbapenem AMR in 19/33 (58%) (85) and 39/117 (33%) (86) clinical isolates. However, another study showed that *in vitro oprD* downregulation in carbapenem-resistant isolates is a rare event, occurring in just 5/40 (13%) isolates (34). As our study illustrates, the markedly different *oprD* expression profiles observed among studies may reflect differing *in vitro* growth conditions; nevertheless, these *in vitro* findings show that *P. aeruginosa* can modulate its *oprD* expression to confer carbapenem AMR. Like *ampC*, our *oprD* results indicate that there may be great clinical value in characterising *in vivo oprD* expression to rapidly identify *P. aeruginosa* infections that are recalcitrant to carbapenem therapy.

### Concluding remarks

Our results add further weight to growing concerns about the relevance of *in vitro* AMR research, particularly *in vitro* gene expression analysis, in clinical practice (87, 88). Due to poor correlation between *in vitro* AMR testing and treatment outcomes in CF, empirical antibiotic selection remains the preferred option due to better patient responses (80). Our findings confirm that alternative approaches must be explored that minimise the transcriptional biases introduced during culture (89) to better reflect *in vivo P. aeruginosa* gene expression (87). We recommend that, wherever possible, gene expression of clinical specimens (e.g. sputum) be tested alongside cultured isolates (20). This information will greatly improve our understanding of the shortcomings of *in vitro* AMR testing whilst aiding in clinical decision making, providing more targeted treatment strategies, enhancing antimicrobial stewardship, and improving patient outcomes. Finally, implementation of microbial-enriched metagenomic sequencing of clinical specimens and *in silico* mixture analysis would permit the detection of AMR determinants within a heterogeneous strain population, providing a superior method to culture-based methods for comprehensive AMR determinant identification.

## Supporting information

Supplementary data for Madden et al., 2022

## Data Availability

All data produced in the present work are contained in the manuscript. Additional data is available upon reasonable request to the authors

## Acknowledgements

This work was supported by Advance Queensland (awards AQIRF0362018 and AQRF13016-17RD2), an Australian Government Research Training Program Scholarship, the Wishlist Sunshine Coast Health Foundation (award 2019-14), the IMPACT Philanthropy Application Program (IPAP2016/01112), UQ-QIMRB (Australian Infectious Disease Grant initiative), the National Health and Medical Research Council (award 455919), The Children’s Health Foundation Queensland (50 0 07), and The Health Innovation, Investment and Research Office of Queensland Health. The funders had no role in study design, data acquisition, analysis, interpretation, writing or submission of the manuscript.

## References

1. Alanis AJ. 2005. Resistance to antibiotics: Are we in the post-antibiotic era? Arch Med Res 36:697–705.

2. Zhen X, Lundborg CS, Sun X, Hu X, Dong H. 2019. Economic burden of antibiotic resistance in ESKAPE organisms: a systematic review. Antimicrob Resist Infect Control 8:137.

3. Mulani MS, Kamble EE, Kumkar SN, Tawre MS, Pardesi KR. 2019. Emerging strategies to combat ESKAPE pathogens in the era of antimicrobial resistance: A review. Front Microbiol 10:539–539.

4. Bassetti M, Vena A, Croxatto A, Righi E, Guery B. 2018. How to manage Pseudomonas aeruginosa infections. Drugs Context 7:1–18.

5. Botelho J, Grosso F, Peixe L. 2019. Antibiotic resistance in Pseudomonas aeruginosa -Mechanisms, epidemiology and evolution. Drug Resist Updat 44:26–47.

6. Breidenstein EB, de la Fuente-Nunez C, Hancock RE. 2011. Pseudomonas aeruginosa: all roads lead to resistance. Trends Microbiol 19:419–26.

7. Shallcross LJ, Howard SJ, Fowler T, Davies SC. 2015. Tackling the threat of antimicrobial resistance: From policy to sustainable action. Philos Trans R Soc Lond B Biol Sci 370:1–5.

8. O’Neill J. 2014. Antimicrobial resistance: Tackling a crisis for the health and wealth of nations. Accessed 12 August 2021.

9. Patel A. 2021. Tackling antimicrobial resistance in the shadow of COVID-19. mBio 12:1-4.

10. Laxminarayan R, Duse A, Wattal C, Zaidi AKM, Wertheim HFL, Sumpradit N, Vlieghe E, Hara GL, Gould IM, Goossens H, Greko C, So AD, Bigdeli M, Tomson G, Woodhouse W, Ombaka E, Peralta AQ, Qamar FN, Mir F, Kariuki S, Bhutta ZA, Coates A, Bergstrom R, Wright GD, Brown ED, Cars O. 2013. Antibiotic resistance - the need for global solutions. Lancet Infect Dis 13:1057–1098.

11. Su M, Satola SW, Read TD. 2019. Genome-based prediction of bacterial antibiotic resistance. J Clin Microbiol 57:1–15.

12. Rossen JWA, Friedrich AW, Moran-Gilad J. 2018. Practical issues in implementing whole-genome-sequencing in routine diagnostic microbiology. Clin Microbiol Infect 24:355–360.

13. Madden DE, McCarthy KL, Bell SC, Olagoke O, Baird T, Neill J, Ramsay KA, Kidd TJ, Stewart AG, Subedi S, Choong K, Fraser TA, Sarovich DS, Price EP. 2021. Rapid fluoroquinolone resistance detection in Pseudomonas aeruginosa using mismatch amplification mutation assay-based real-time PCR. medRxiv.

14. Richardot C, Plesiat P, Fournier D, Monlezun L, Broutin I, Llanes C. 2015. Carbapenem resistance in cystic fibrosis strains of Pseudomonas aeruginosa as a result of amino acid substitutions in porin OprD. Int J Antimicrob Agents 45:529–32.

15. Godfroid M, Dagan T, Merker M, Kohl TA, Diel R, Maurer FP, Niemann S, Kupczok A. 2020. Insertion and deletion evolution reflects antibiotics selection pressure in a Mycobacterium tuberculosis outbreak. PLoS Pathog 16:1–24.

16. Huang TW, Chen TL, Chen YT, Lauderdale TL, Liao TL, Lee YT, Chen CP, Liu YM, Lin AC, Chang YH, Wu KM, Kirby R, Lai JF, Tan MC, Siu LK, Chang CM, Fung CP, Tsai SF. 2013. Copy number change of the NDM-1 sequence in a multidrug-resistant Klebsiella pneumoniae clinical isolate. PLoS One 8:1–12.

17. Chalhoub H, Sáenz Y, Rodriguez-Villalobos H, Denis O, Kahl BC, Tulkens PM, Van Bambeke F. 2016. High-level resistance to meropenem in clinical isolates of Pseudomonas aeruginosa in the absence of carbapenemases: role of active efflux and porin alterations. Int J Antimicrob Agents 48:740–743.

18. Stewart AG, Price EP, Schabacker K, Birikmen M, Harris PNA, Choong K, Subedi S, Sarovich DS. 2021. Molecular epidemiology of third-generation cephalosporin-resistant Enterobacteriaceae in Southeast Queensland, Australia. Antimicrob Agents Chemother 65:1–13.

19. Chantratita N, Rholl DA, Sim B, Wuthiekanun V, Limmathurotsakul D, Amornchai P, Thanwisai A, Chua HH, Ooi WF, Holden MT, Day NP, Tan P, Schweizer HP, Peacock SJ. 2011. Antimicrobial resistance to ceftazidime involving loss of penicillin-binding protein 3 in Burkholderia pseudomallei. Proc Natl Acad Sci U S A 108:17165–70.

20. Martin LW, Robson CL, Watts AM, Gray AR, Wainwright CE, Bell SC, Ramsay KA, Kidd TJ, Reid DW, Brockway B, Lamont IL. 2018. Expression of Pseudomonas aeruginosa antibiotic resistance genes varies greatly during infections in cystic fibrosis patients. Antimicrob Agents Chemother 62:1–11.

21. Wheatley R, Diaz Caballero J, Kapel N, de Winter Fhr, Jangir P, Quinn A, del Barrio-Tofiño E, López-Causapé C, Hedge J, Torrens G, Van der Schalk T, Xavier BB, Fernández-Cuenca F, Arenzana A, Recanatini C, Timbermont L, Sifakis F, Ruzin A, Ali O, Lammens C, Goossens H, Kluytmans J, Kumar-Singh S, Oliver A, Malhotra-Kumar S, MacLean C. 2021. Rapid evolution and host immunity drive the rise and fall of carbapenem resistance during an acute Pseudomonas aeruginosa infection. Nat Commun 12:1–12.

22. Lee JY, Ko KS. 2012. OprD mutations and inactivation, expression of efflux pumps and AmpC, and metallo-beta-lactamases in carbapenem-resistant Pseudomonas aeruginosa isolates from South Korea. Int J Antimicrob Agents 40:168–72.

23. Cabot G, Ocampo-Sosa AA, Tubau F, Macia MD, Rodriguez C, Moya B, Zamorano L, Suarez C, Pena C, Martinez-Martinez L, Oliver A. 2011. Overexpression of AmpC and efflux pumps in Pseudomonas aeruginosa isolates from bloodstream infections: prevalence and impact on resistance in a Spanish multicenter study. Antimicrob Agents Chemother 55:1906–11.

24. Webb JR, Price EP, Somprasong N, Schweizer HP, Baird RW, Currie BJ, Sarovich DS. 2018. Development and validation of a triplex quantitative real-time PCR assay to detect efflux pump-mediated antibiotic resistance in Burkholderia pseudomallei. Future Microbiol 12:1403–1418.

25. Poole K. 2011. Pseudomonas aeruginosa: Resistance to the max. Front Microbiol 2:1–13.

26. Juan C, Moya B, Perez JL, Oliver A. 2006. Stepwise upregulation of the Pseudomonas aeruginosa chromosomal cephalosporinase conferring high-level beta-lactam resistance involves three AmpD homologues. Antimicrob Agents Chemother 50:1780–7.

27. Shu JC, Kuo AJ, Su LH, Liu TP, Lee MH, Su IN, Wu TL. 2017. Development of carbapenem resistance in Pseudomonas aeruginosa is associated with OprD polymorphisms, particularly the amino acid substitution at codon 170. J Antimicrob Chemother 72:2489–2495.

28. Sherrard LJ, Wee BA, Duplancic C, Ramsay KA, Dave KA, Ballard E, Wainwright CE, Grimwood K, Sidjabat HE, Whiley DM, Beatson SA, Kidd TJ, Bell SC. 2021. Emergence and impact of oprD mutations in Pseudomonas aeruginosa strains in cystic fibrosis. J Cyst Fibros 21:1–9.

29. Lee JK, Lee YS, Park YK, Kim BS. 2005. Alterations in the GyrA and GyrB subunits of topoisomerase II and the ParC and ParE subunits of topoisomerase IV in ciprofloxacin-resistant clinical isolates of Pseudomonas aeruginosa. Int J Antimicrob Agents 25:290–5.

30. Poole K, Tetro K, Zhao Q, Neshat S, Heinrichs DE, Bianco N. 1996. Expression of the multidrug resistance operon mexA-mexB-oprM in Pseudomonas aeruginosa: mexR encodes a regulator of operon expression. Antimicrob Agents Chemother 40:2021–8.

31. Chen W, Wang D, Zhou W, Sang H, Liu X, Ge Z, Zhang J, Lan L, Yang CG, Chen H. 2016. Novobiocin binding to NalD induces the expression of the MexAB-OprM pump in Pseudomonas aeruginosa. Mol Microbiol 100:749–58.

32. Köhler T, Epp SF, Curty LK, Pechère JC. 1999. Characterization of MexT, the regulator of the MexE-MexF-OprN multidrug efflux system of Pseudomonas aeruginosa. J Bacteriol 181:6300–5.

33. Morita Y, Tomida J, Kawamura Y. 2012. MexXY multidrug efflux system of Pseudomonas aeruginosa. Front Microbiol 3:1–13.

34. Aghazadeh M, Hojabri Z, Mahdian R, Nahaei MR, Rahmati M, Hojabri T, Pirzadeh T, Pajand O. 2014. Role of efflux pumps: MexAB-OprM and MexXY(-OprA), AmpC cephalosporinase and OprD porin in non-metallo-beta-lactamase producing Pseudomonas aeruginosa isolated from cystic fibrosis and burn patients. Infect Genet Evol 24:187–92.

35. Poole K, Gotoh N, Tsujimoto H, Zhao Q, Wada A, Yamasaki T, Neshat S, Yamagishi J, Li XZ, Nishino T. 1996. Overexpression of the mexC-mexD-oprJ efflux operon in nfxB-type multidrug-resistant strains of Pseudomonas aeruginosa. Mol Microbiol 21:713–24.

36. Fraud S, Campigotto AJ, Chen Z, Poole K. 2008. MexCD-OprJ multidrug efflux system of Pseudomonas aeruginosa: Involvement in chlorhexidine resistance and induction by membrane-damaging agents dependent upon the AlgU stress response sigma factor. Antimicrob Agents Chemother 52:4478–4482.

37. Gotoh N, Tsujimoto H, Tsuda M, Okamoto K, Nomura A, Wada T, Nakahashi M, Nishino T. 1998. Characterization of the MexC-MexD-OprJ Multidrug Efflux System in mexA-mexB-oprM Mutants of Pseudomonas aeruginosa. Antimicrob Agents Chemother 42:1938–1943.

38. Oh H, Stenhoff J, Jalal S, Wretlind B. 2003. Role of efflux pumps and mutations in genes for topoisomerases II and IV in fluoroquinolone-resistant Pseudomonas aeruginosa strains. Microb Drug Resist 9:323–8.

39. Islam S, Jalal S, Wretlind B. 2004. Expression of the MexXY efflux pump in amikacin-resistant isolates of Pseudomonas aeruginosa. Clin Microbiol Infect 10:877–83.

40. Webb KA, Olagoke O, Baird T, Neill J, Pham A, Wells TJ, Ramsay KA, Bell SC, Sarovich DS, Price EP. 2021. Genomic diversity and antimicrobial resistance of Prevotella spp. isolated from chronic lung disease airways. Microb Genom 8:000754.

41. Nadkarni MA, Martin FE, Jacques NA, Hunter N. 2002. Determination of bacterial load by real-time PCR using a broad-range (universal) probe and primers set. Microbiology (Reading) 148:257–266.

42. de Lamballerie X, Zandotti C, Vignoli C, Bollet C, de Micco P. 1992. A one-step microbial DNA extraction method using “Chelex 100” suitable for gene amplification. Res Microbiol 143:785–90.

43. Anuj SN, Whiley DM, Kidd TJ, Bell SC, Wainwright CE, Nissen MD, Sloots TP. 2009. Identification of Pseudomonas aeruginosa by a duplex real-time polymerase chain reaction assay targeting the ecfX and the gyrB genes. Diagn Microbiol Infect Dis 63:127–31.

44. Horcajada JP, Montero M, Oliver A, Sorlí L, Luque S, Gómez-Zorrilla S, Benito N, Grau S. 2019. Epidemiology and treatment of multidrug-resistant and extensively drug-resistant Pseudomonas aeruginosa infections. Clin Microbiol Rev 32:1–52.

45. Masuda N, Sakagawa E, Ohya S, Gotoh N, Tsujimoto H, Nishino T. 2000. Substrate specificities of MexAB-OprM, MexCD-OprJ, and MexXY-OprM efflux pumps in Pseudomonas aeruginosa. Antimicrob Agents Chemother 44:3322–7.

46. Buffet-Bataillon S, Tattevin P, Maillard J-Y, Bonnaure-Mallet M, Jolivet-Gougeon A. 2016. Efflux pump induction by quaternary ammonium compounds and fluoroquinolone resistance in bacteria. Future Microbiol 11:81–92.

47. Fetar H, Gilmour C, Klinoski R, Daigle DM, Dean CR, Poole K. 2011. mexEF-oprN multidrug efflux operon of Pseudomonas aeruginosa: Regulation by the MexT activator in response to nitrosative stress and chloramphenicol. Antimicrob Agents Chemother 55:508–514.

48. Dunne WM, Jr., Hardin DJ. 2005. Use of several inducer and substrate antibiotic combinations in a disk approximation assay format to screen for AmpC induction in patient isolates of Pseudomonas aeruginosa, Enterobacter spp., Citrobacter spp., and Serratia spp. J of Clin Microbiol 43:5945–5949.

49. Oliver A, Baquero F, Blázquez J. 2002. The mismatch repair system (mutS, mutL and uvrD genes) in Pseudomonas aeruginosa: Molecular characterization of naturally occurring mutants. Mol Microbiol 43:1641–1650.

50. van Belkum A, Soriaga LB, LaFave MC, Akella S, Veyrieras J-B, Barbu EM, Shortridge D, Blanc B, Hannum G, Zambardi G, Miller K, Enright MC, Mugnier N, Brami D, Schicklin S, Felderman M, Schwartz AS, Richardson TH, Peterson TC, Hubby B, Cady KC. 2015. Phylogenetic distribution of CRISPR-Cas systems in antibiotic-resistant Pseudomonas aeruginosa. mBio 6:1–13.

51. Cabot G, López-Causapé C, Ocampo-Sosa AA, Sommer LM, Domínguez MÁ, Zamorano L, Juan C, Tubau F, Rodríguez C, Moyà B, Peña C, Martínez-Martínez L, Plesiat P, Oliver A. 2016. Deciphering the resistome of the widespread Pseudomonas aeruginosa sequence type 175 international high-risk clone through whole-genome sequencing. Antimicrob Agents Chemother 60:7415–7423.

52. Kos VN, Déraspe M, McLaughlin RE, Whiteaker JD, Roy PH, Alm RA, Corbeil J, Gardner H. 2015. The resistome of Pseudomonas aeruginosa in relationship to phenotypic susceptibility. Antimicrob Agents Chemother 59:427–436.

53. Del Barrio-Tofiño E, López-Causapé C, Cabot G, Rivera A, Benito N, Segura C, Montero MM, Sorlí L, Tubau F, Gómez-Zorrilla S, Tormo N, Durá-Navarro R, Viedma E, Resino-Foz E, Fernández-Martínez M, González-Rico C, Alejo-Cancho I, Martínez JA, Labayru-Echverria C, Dueñas C, Ayestarán I, Zamorano L, Martinez-Martinez L, Horcajada JP, Oliver A. 2017. Genomics and susceptibility profiles of extensively drug-resistant Pseudomonas aeruginosa isolates from Spain. Antimicrob Agents Chemother 61:1–13.

54. Sherrard LJ, Tai AS, Wee BA, Ramsay KA, Kidd TJ, Ben Zakour NL, Whiley DM, Beatson SA, Bell SC. 2017. Within-host whole genome analysis of an antibiotic resistant Pseudomonas aeruginosa strain sub-type in cystic fibrosis. PLoS ONE 12:1–15.

55. Fraser TA, Bell MG, Harris PNA, Bell SC, Bergh H, Nguyen T-K, Kidd TJ, Nimmo GR, Sarovich DS, Price EP. 2019. Quantitative real-time PCR assay for the rapid identification of the intrinsically multidrug-resistant bacterial pathogen Stenotrophomonas maltophilia. Microb Genom 5:2–11.

56. Price EP, Dale JL, Cook JM, Sarovich DS, Seymour ML, Ginther JL, Kaufman EL, Beckstrom-Sternberg SM, Mayo M, Kaestli M, Glass MB, Gee JE, Wuthiekanun V, Warner JM, Baker A, Foster JT, Tan P, Tuanyok A, Limmathurotsakul D, Peacock SJ, Currie BJ, Wagner DM, Keim P, Pearson T. 2012. Development and validation of Burkholderia pseudomallei-specific real-time PCR assays for clinical, environmental or forensic detection applications. PLoS One 7:1–9.

57. Araujo P. 2009. Key aspects of analytical method validation and linearity evaluation. J Chromatogr B Analyt Technol Biomed Life Sci 877:2224–34.

58. Livak KJ, Schmittgen TD. 2001. Analysis of relative gene expression data using real-time quantitative PCR and the 2(-Delta Delta C(T)) Method. Methods 25:402–8.

59. Madden DE, Webb JR, Steinig EJ, Currie BJ, Price EP, Sarovich DS. 2021. Taking the next-gen step: Comprehensive antimicrobial resistance detection from Burkholderia pseudomallei. EBioMedicine 63:103152.

60. Wood DE, Lu J, Langmead B. 2019. Improved metagenomic analysis with Kraken 2. Genome Biol 20:257.

61. Pedersen BS, Quinlan AR. 2018. Mosdepth: quick coverage calculation for genomes and exomes. Bioinformatics 34:867–868.

62. Bortolaia V, Kaas RS, Ruppe E, Roberts MC, Schwarz S, Cattoir V, Philippon A, Allesoe RL, Rebelo AR, Florensa AF, Fagelhauer L, Chakraborty T, Neumann B, Werner G, Bender JK, Stingl K, Nguyen M, Coppens J, Xavier BB, Malhotra-Kumar S, Westh H, Pinholt M, Anjum MF, Duggett NA, Kempf I, Nykäsenoja S, Olkkola S, Wieczorek K, Amaro A, Clemente L, Mossong J, Losch S, Ragimbeau C, Lund O, Aarestrup FM. 2020. ResFinder 4.0 for predictions of phenotypes from genotypes. Journal of Antimicrobial Chemotherapy 75:3491–3500.

63. Juan C, Macia MD, Gutierrez O, Vidal C, Perez JL, Oliver A. 2005. Molecular mechanisms of β-lactam resistance mediated by AmpC hyperproduction in Pseudomonas aeruginosa clinical strains. Antimicrob Agents Chemother 49:4733–8.

64. Kong KF, Aguila A, Schneper L, Mathee K. 2010. Pseudomonas aeruginosa beta-lactamase induction requires two permeases, AmpG and AmpP. BMC Microbiol 10:328.

65. Tomás M, Doumith M, Warner M, Turton JF, Beceiro A, Bou G, Livermore DM, Woodford N. 2010. Efflux pumps, OprD porin, AmpC beta-lactamase, and multiresistance in Pseudomonas aeruginosa isolates from cystic fibrosis patients. Antimicrob Agents Chemother 54:2219–24.

66. Purssell A, Poole K. 2013. Functional characterization of the NfxB repressor of the mexCD-oprJ multidrug efflux operon of Pseudomonas aeruginosa. Microbiology (Reading) 159:2058–2073.

67. Livermore DM. 1995. Beta-Lactamases in laboratory and clinical resistance. Clin Microbiol Rev 8:557–84.

68. Rehman A, Jeukens J, Levesque RC, Lamont IL. 2021. Gene-gene interactions dictate ciprofloxacin resistance in Pseudomonas aeruginosa and facilitate prediction of resistance phenotype from genome sequence data. Antimicrob Agents Chemother 65:1–15.

69. Clark ST, Sinha U, Zhang Y, Wang PW, Donaldson SL, Coburn B, Waters VJ, Yau YCW, Tullis DE, Guttman DS, Hwang DM. 2019. Penicillin-binding protein 3 is a common adaptive target among Pseudomonas aeruginosa isolates from adult cystic fibrosis patients treated with β-lactams. Int J Antimicrob Agents 53:620–628.

70. Bolard A, Plésiat P, Jeannot K. 2018. Mutations in gene fusA1 as a novel mechanism of aminoglycoside resistance in clinical strains of Pseudomonas aeruginosa. Antimicrob Agents Chemother 62:1–10.

71. Köhler T, Michéa-Hamzehpour M, Henze U, Gotoh N, Curty LK, Pechère JC. 1997. Characterization of MexE-MexF-OprN, a positively regulated multidrug efflux system of Pseudomonas aeruginosa. Mol Microbiol 23:345–54.

72. Feng X, Zhang Z, Li X, Song Y, Kang J, Yin D, Gao Y, Shi N, Duan J. 2019. Mutations in gyrB play an important role in ciprofloxacin-resistant Pseudomonas aeruginosa. Infect Drug Resist 12:261–272.

73. Higgins PG, Fluit AC, Milatovic D, Verhoef J, Schmitz FJ. 2003. Mutations in GyrA, ParC, MexR and NfxB in clinical isolates of Pseudomonas aeruginosa. Int J Antimicrob Agents 21:409–13.

74. Dötsch A, Becker T, Pommerenke C, Magnowska Z, Jänsch L, Häussler S. 2009. Genomewide identification of genetic determinants of antimicrobial drug resistance in Pseudomonas aeruginosa. Antimicrob Agents Chemother 53:2522–31.

75. Colque CA, Albarracin Orio AG, Feliziani S, Marvig RL, Tobares AR, Johansen HK, Molin S, Smania AM. 2020. Hypermutator Pseudomonas aeruginosa Exploits Multiple Genetic Pathways To Develop Multidrug Resistance during Long-Term Infections in the Airways of Cystic Fibrosis Patients. Antimicrob Agents Chemother 64.

76. Zamorano L, Moya B, Juan C, Oliver A. 2010. Differential beta-lactam resistance response driven by ampD or dacB (PBP4) inactivation in genetically diverse Pseudomonas aeruginosa strains. J Antimicrob Chemother 65:1540–2.

77. Winstanley C, O’Brien S, Brockhurst MA. 2016. Pseudomonas aeruginosa evolutionary adaptation and diversification in cystic fibrosis chronic lung infections. Trends Microbiol 24:327–337.

78. Valderrey AD, Pozuelo MJ, Jiménez PA, Maciá MD, Oliver A, Rotger R. 2010. Chronic colonization by Pseudomonas aeruginosa of patients with obstructive lung diseases: cystic fibrosis, bronchiectasis, and chronic obstructive pulmonary disease. Diagn Microbiol Infect Dis 68:20–7.

79. Lee M, Hesek D, Blázquez B, Lastochkin E, Boggess B, Fisher JF, Mobashery S. 2015. Catalytic spectrum of the penicillin-binding protein 4 of Pseudomonas aeruginosa, a nexus for the induction of β-lactam antibiotic resistance. J Am Chem Soc 137:190–200.

80. Van den Bossche S, De Broe E, Coenye T, Van Braeckel E, Crabbé A. 2021. The cystic fibrosis lung microenvironment alters antibiotic activity: Causes and effects. Eur Respir Rev 30:1–16.

81. Vettoretti L, Plesiat P, Muller C, El Garch F, Phan G, Attree I, Ducruix A, Llanes C. 2009. Efflux unbalance in Pseudomonas aeruginosa isolates from cystic fibrosis patients. Antimicrob Agents Chemother 53:1987–97.

82. Goodman AL, Kulasekara B, Rietsch A, Boyd D, Smith RS, Lory S. 2004. A signaling network reciprocally regulates genes associated with acute infection and chronic persistence in Pseudomonas aeruginosa. Dev Cell 7:745–54.

83. Werner E, Roe F, Bugnicourt A, Franklin MJ, Heydorn A, Molin S, Pitts B, Stewart PS. 2004. Stratified growth in Pseudomonas aeruginosa biofilms. Appl Environ Microbiol 70:6188–96.

84. Murphy TF. 2008. The many faces of Pseudomonas aeruginosa in chronic obstructive pulmonary disease. Clin Infect Dis 47:1534–1536.

85. Quale J, Bratu S, Gupta J, Landman D. 2006. Interplay of efflux system, ampC, and oprD expression in carbapenem resistance of Pseudomonas aeruginosa clinical isolates. Antimicrob Agents Chemother 50:1633–1641.

86. Pai H, Kim J, Kim J, Lee JH, Choe KW, Gotoh N. 2001. Carbapenem resistance mechanisms in Pseudomonas aeruginosa clinical isolates. Antimicrob Agents Chemother 45:480–4.

87. Fung C, Naughton S, Turnbull L, Tingpej P, Rose B, Arthur J, Hu H, Harmer C, Harbour C, Hassett DJ, Whitchurch CB, Manos J. 2010. Gene expression of Pseudomonas aeruginosa in a mucin-containing synthetic growth medium mimicking cystic fibrosis lung sputum. J Med Microbiol 59:1089–1100.

88. Hill D, Rose B, Pajkos A, Robinson M, Bye P, Bell S, Elkins M, Thompson B, MacLeod C, Aaron SD, Harbour C. 2005. Antibiotic susceptibilities of Pseudomonas aeruginosa isolates derived from patients with cystic fibrosis under aerobic, anaerobic, and biofilm conditions. J of Clin Microbiol 43:5085–5090.

89. Kirchner S, Fothergill JL, Wright EA, James CE, Mowat E, Winstanley C. 2012. Use of artificial sputum medium to test antibiotic efficacy against Pseudomonas aeruginosa in conditions more relevant to the cystic fibrosis lung. J Vis Exp 64:1–7.

