## Supplementary data for Madden et al., 2022 for "Express yourself: Quantitative real-time PCR assays for rapid chromosomal antimicrobial resistance detection in *Pseudomonas aeruginosa*"

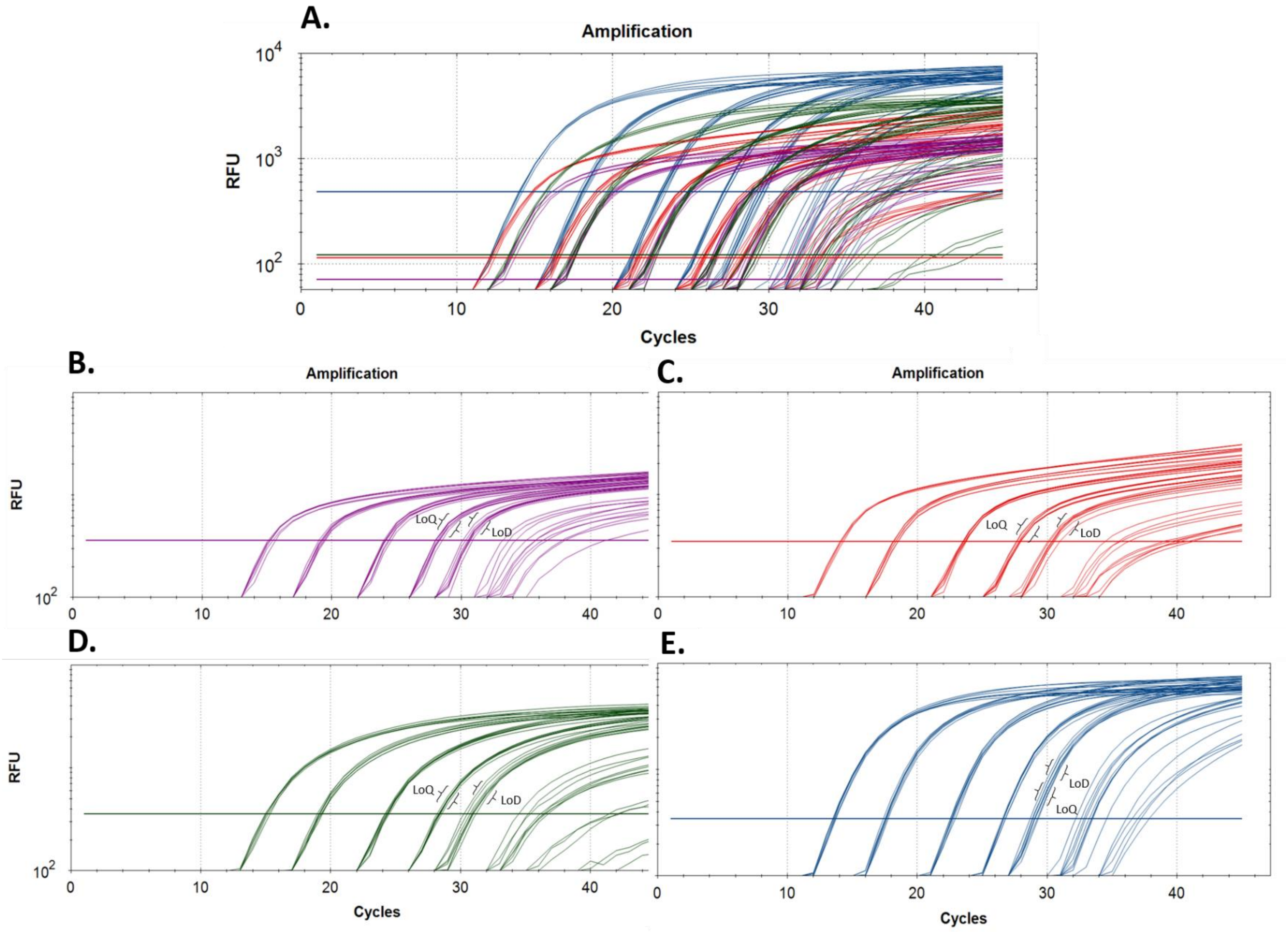


**Figure S1. A.** Quantitative PCR amplification plots for *Pseudomonas aeruginosa* multiplex #1 (quadriplex) assay. **B.** *rpsL*, **C.** *ampC*, **D.** *mexY* and **E.** *mexB*. Limits of detection and quantification were determined as described in Methods section 'qPCR assay limits of detection (LoD) and quantification (LoQ)'.

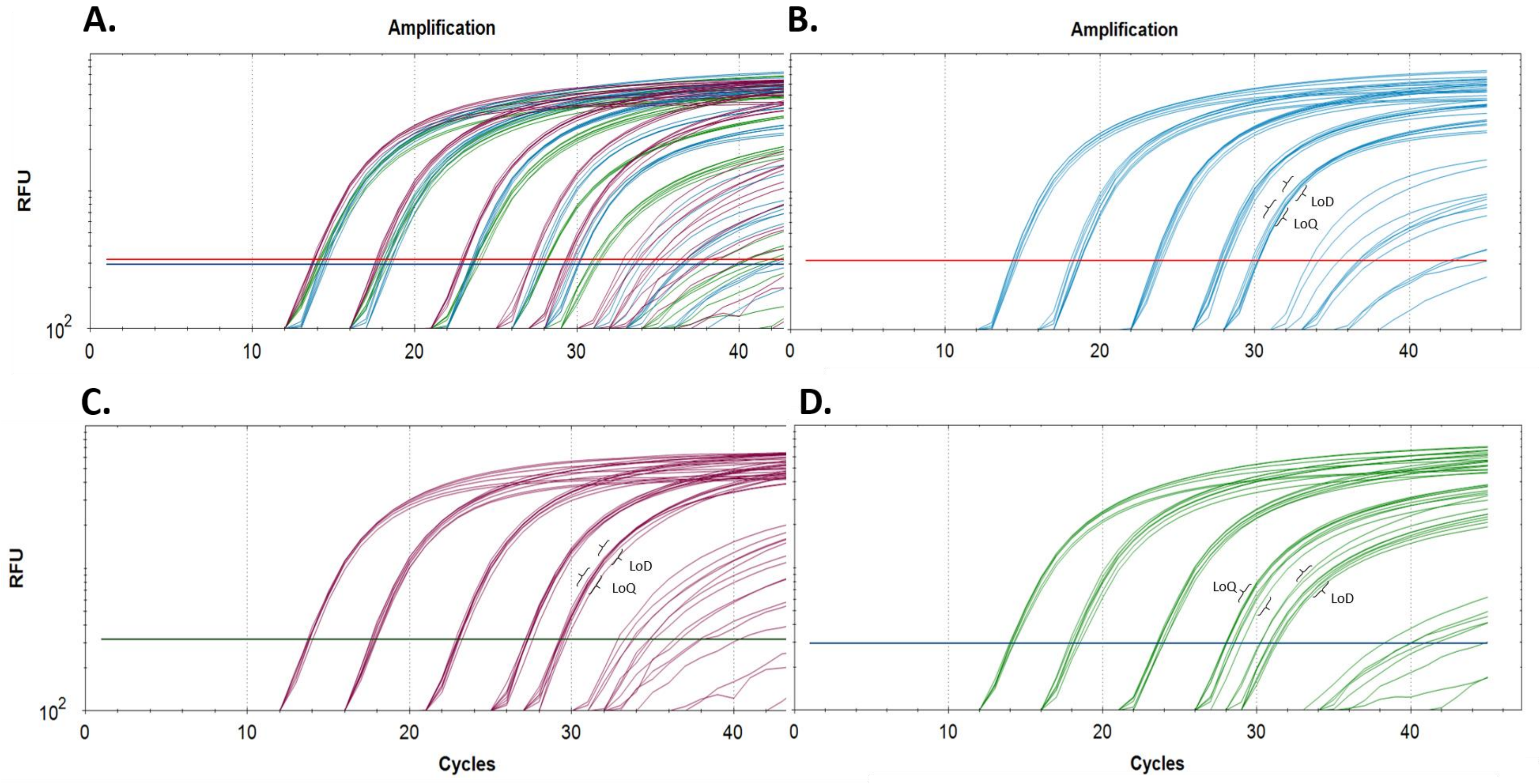


**Figure S2. A.** Quantitative PCR amplification plots for *Pseudomonas aeruginosa* multiplex #2 (triplex) assay. **B.** *mexE*, **C.** *mexC* and **D.** *oprD.* Limits of detection and quantification were determined as described in Methods section 'qPCR assay limits of detection (LoD) and quantification (LoQ)'.

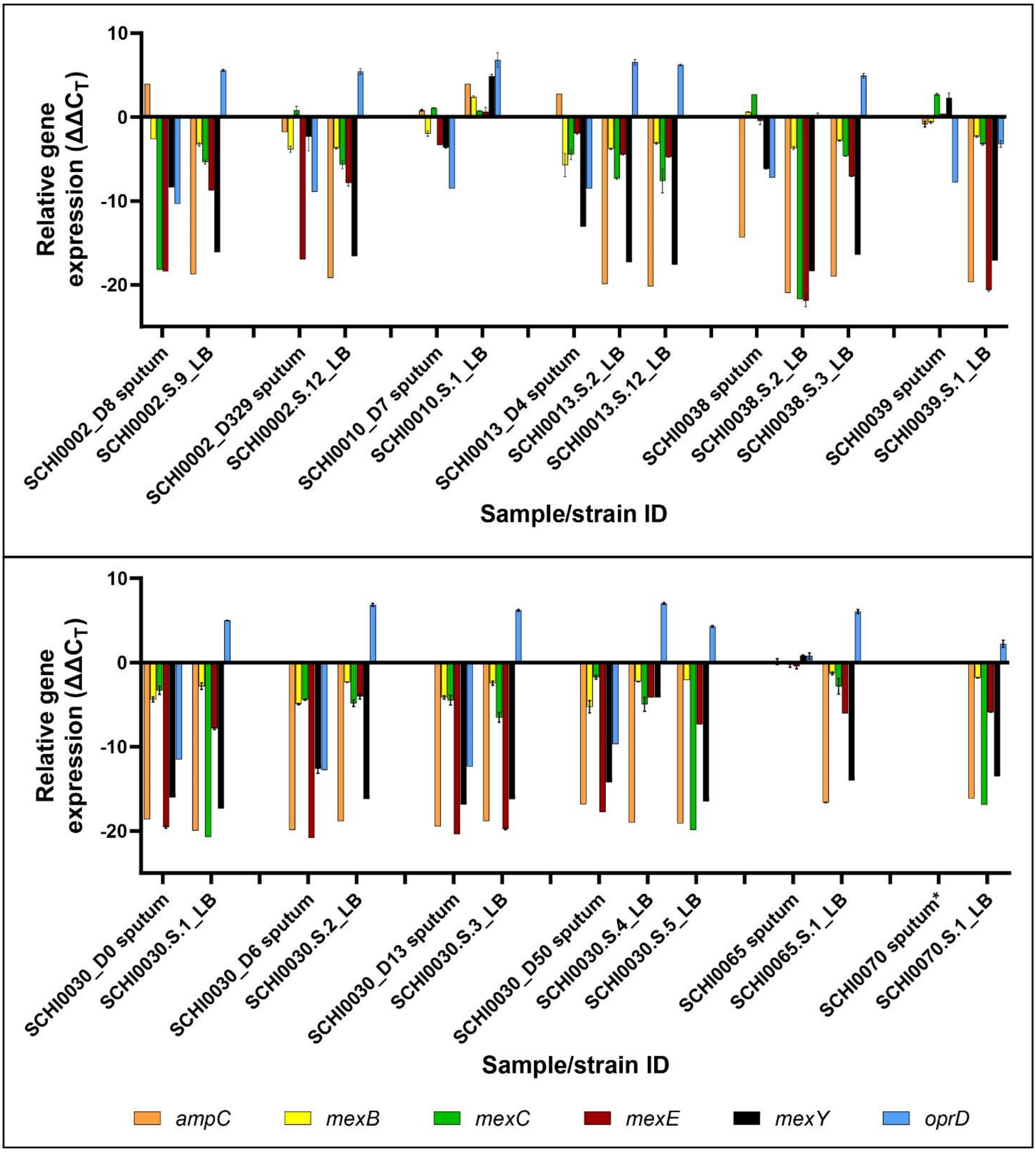


**Figure S3.** Differential expression (relative $\log_2$ fold change) of *Pseudomonas aeruginosa* in sputa and their derived cultures (denoted by '.S.') when grown in Luria-Bertani broth. Gene expression for each locus was first normalised against *rpsL* ($\Delta C_T$); relative expression was then determined by comparing $\Delta C_T$ values against sputum from COPD participant SCHI0070 (i.e. $\Delta\Delta C_T$), which is denoted by an asterisk.

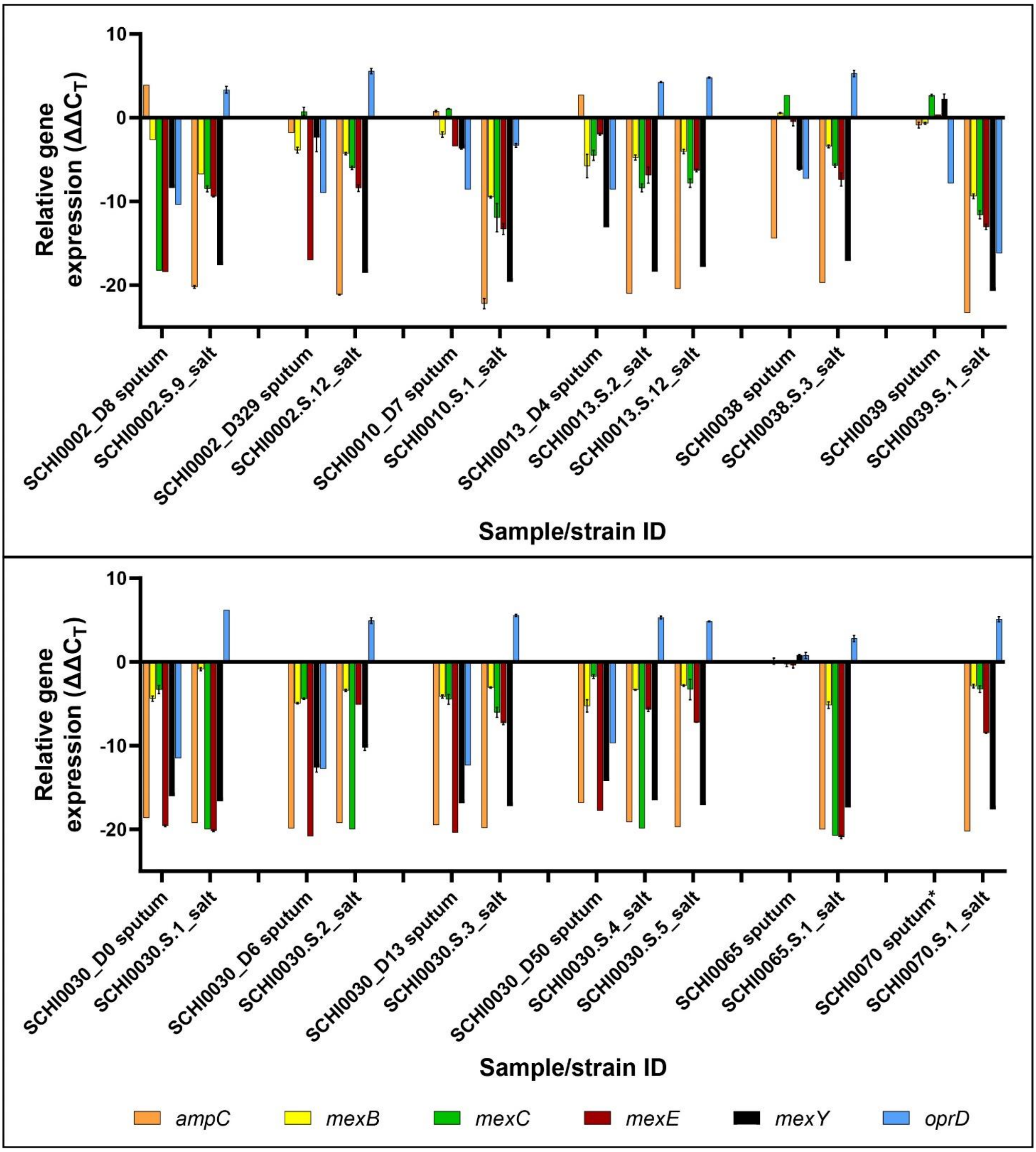


**Figure S4.** Differential expression (relative $\log_2$ fold change) of *Pseudomonas aeruginosa* in sputa and their derived cultures (denoted by '.S.') when grown in Luria-Bertani broth containing an additional 2.5% NaCl ('salt'). Gene expression for each locus was first normalised against *rpsL* ($\Delta C_T$); relative expression was then determined by comparing $\Delta C_T$ values against sputum from COPD participant SCHI0070 (i.e. $\Delta\Delta C_T$), which is denoted by an asterisk.

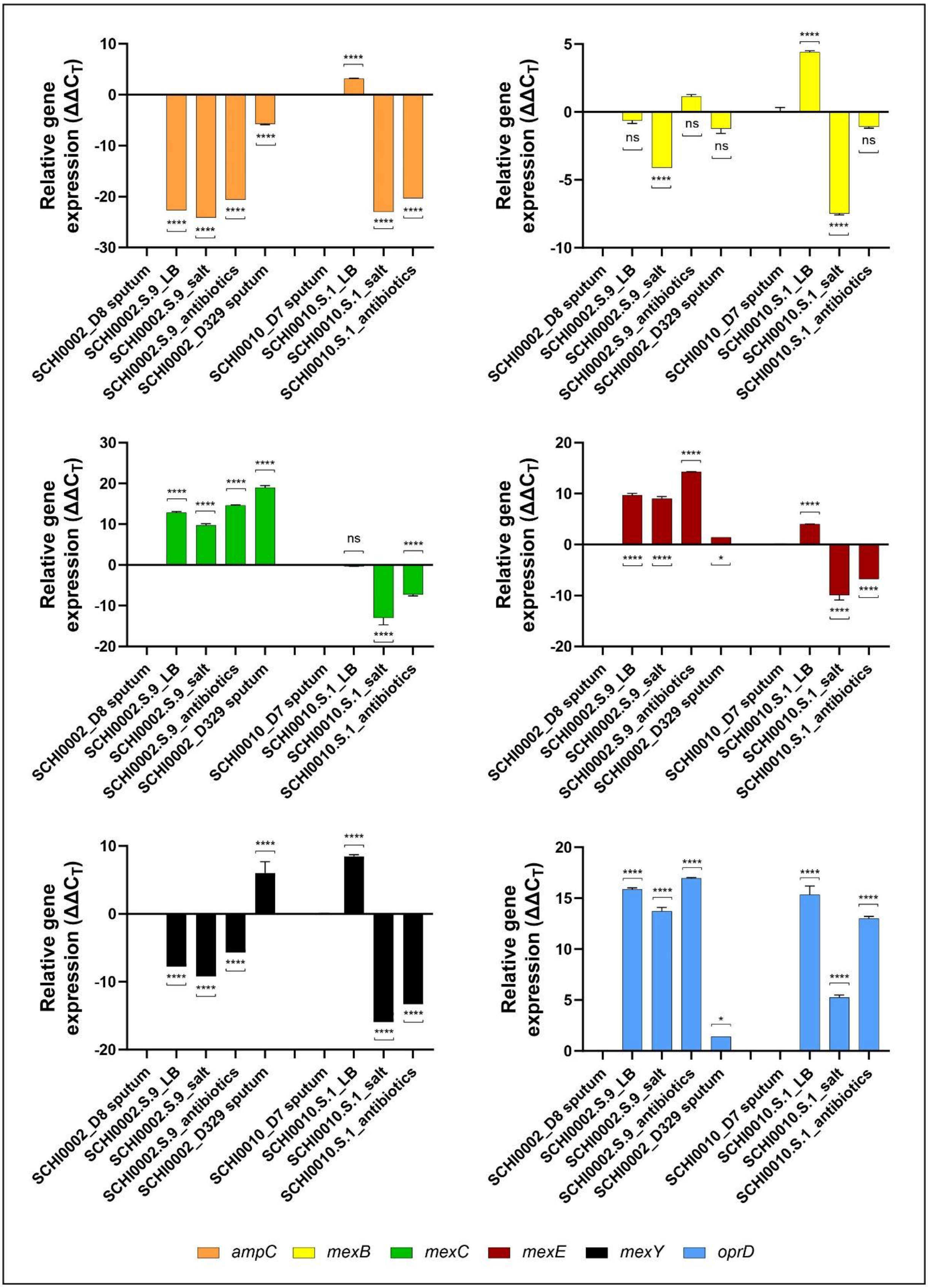


**Figure S5.** Differential expression (relative $\log_2$ fold change) of *Pseudomonas aeruginosa* loci in cystic fibrosis SCHI0002 and SCHI0010 sputa compared with derived cultures grown in Luria-Bertani broth ('LB'),

LB broth plus 2.5% NaCl (‘salt’), and LB broth containing a 5-antibiotic cocktail (‘antibiotics’). Gene expression for each locus was first normalised against *rpsL* ($\Delta C_T$); relative expression was then determined by comparing $\Delta C_T$ values against COPD participant SCHI0070, who was not receiving anti-pseudomonal antibiotics at the time of sputum collection ($\Delta\Delta C_T$). Significance was calculated using 2-way ANOVA analysis and Tukey’s multiple comparison test when compared with either SCHI0002_D8 or SCHI0010_D7 sputa. *, $p$ <0.05; ***, $p$ <0.001; ****, $p$ <0.0001; ns, not significant.

**Table S1. *Pseudomonas aeruginosa rpsL* housekeeping gene expression in all tested cystic fibrosis (CF) and chronic obstructive pulmonary disease (COPD) sputa.** All samples were tested in duplicate. Samples in black text had moderate to high *P. aeruginosa* load according to culture; samples in orange text had low *P. aeruginosa* load according to culture; samples in red text were culture-negative for *P. aeruginosa* and included as negative controls. The orange and red text samples were excluded from further analysis due to *rpsL* cycles-to-threshold ($C_T$) values exceeding the limit of quantitation.

| Participant | Specimen type | Airway disease | Average $C_T$ value | Standard deviation |
|---|---|---|---|---|
| SCHI0002_D8 | Sputum | CF | 26.7 | 0.00 |
| SCHI0002_D329 | Sputum | CF | 28.1 | 0.05 |
| SCHI0010_D7 | Sputum | CF | 28.4 | 0.06 |
| SCHI0013_D4 | Sputum | CF | 28.4 | 0.28 |
| SCHI0018_D8 | Sputum | CF | 28.6 | 0.00 |
| SCHI0030_D0 | Sputum | CF | 25.5 | 0.23 |
| SCHI0030_D6 | Sputum | CF | 24.3 | 0.20 |
| SCHI0030_D13 | Sputum | CF | 24.7 | 0.68 |
| SCHI0030_D50 | Sputum | CF | 27.3 | 0.18 |
| SCHI0038 | Sputum | COPD | 29.7 | 0.02 |
| SCHI0039 | Sputum | COPD | 29.2 | 0.38 |
| SCHI0042 | Sputum | COPD | No amplification | --- |
| SCHI0049 | Bronchial washing | COPD | No amplification | --- |
| SCHI0050_D3 | Sputum | COPD | 27.3 | 0.02 |
| SCHI0052_D14 | Sputum | COPD | No amplification | --- |
| SCHI0052_D39 | Sputum | COPD | 32.1 | 0.35 |
| SCHI0054_D0 | Sputum | COPD | 34.2* | --- |
| SCHI0054_D13 | Sputum | COPD | No amplification | --- |
| SCHI0064 | Sputum | COPD | 36.4* | --- |
| SCHI0065 | Sputum | COPD | 25.1 | 0.04 |
| SCHI0067 | Sputum | COPD | No amplification | --- |
| SCHI0070_D2 | Sputum | COPD | 23.0 | 0.01 |
| SCHI0071 | Sputum | COPD | 35.3* | --- |
| SCHI0086 | Sputum | COPD | No amplification | --- |
| SCHI0109 | Sputum | COPD | 22.3 | 0.08 |

*Only 1 of 2 replicates amplified

**Table S2. Number of mixed variant sites identified in *Pseudomonas aeruginosa* from sputum 'culturomes'.** When compared with the number of mixed variants identified in purified, derived isolates (where no mixtures were expected), all tested sputa except for SCHI0002_D8 demonstrated strain homogeneity. The cystic fibrosis (CF) SCHI0002_D8 culturome in particular possessed a considerable heterogeneous *P. aeruginosa* strain population.

| Participant | Sample type | Mixed SNP sites |
|---|---|---|
| SCHI0002_D8 | CF sputum | 21664 |
| SCHI0002.S.8 | Isolate | 1150 |
| SCHI0002.S.9 | Isolate | 1049 |
| SCHI0002_D329 | CF sputum | 833 |
| SCHI0002.S.12 | Isolate | 1003 |
| SCHI0010_D7 | CF sputum | 1521 |
| SCHI0010.S.1 | Isolate | 1769 |
| SCHI0013_D4 | CF sputum | 701 |
| SCHI0013.S.2 | Isolate | 970 |
| SCHI0013.S.12 | Isolate | 1535 |
| SCHI0038 | COPD sputum | 1039 |
| SCHI0038.S.3 | Isolate | 849 |
| SCHI0039 | COPD sputum | 688 |
| SCHI0039.S.1 | Isolate | 760 |
| SCHI0050_D3 | COPD sputum | 678 |
| SCHI0050.S.1 | Isolate | 691 |
